# Clinical Features, Outcomes of Treatments, Inflammasome Function and Longitudinal Clonal Dynamics into *NLRP3* Mosaicism: Evidence from the Largest Cryopyrin-associated Periodic Syndromes Cohort to Date

**DOI:** 10.1101/2024.12.15.24318703

**Authors:** Nuria Bonet, Jose M. Mascaro, Laura Hurtado-Navarro, Diego Angosto-Bazarra, Jose Luis Callejas-Rubio, Daniel Clemente, Alejandro Souto, Olalla Lima, Natalia Palmou-Fontana, Eulalia Baselga, Santiago Jiménez-Treviño, Agustin Remesal, Marta Andreu-Barasoain, Luis Fernandez-Dominguez, Josep Riera-Monroig, Maria Aparicio, Juan Garcia-Herrero, David Pesqué, Maria Teresa Sanchez-Calvin, Jose Miguel Lezana-Rosales, Maria Correyero-Plaza, Julio Garcia-Villalba, Victor Bolaño, Sara Peiro, Mar Diaz, Alexandru Vlagea, Daniel Lorca, Virginia Fabregat, Maria Carmen Anton, Susana Plaza, Luis Ignacio Gonzalez-Granado, Concepción Postigo, Jose Maria Garcia-Ruiz de Morales, Enrique Gómez de la Fuente, Estibaliz Iglesias, Javier Gomez-Roman, Caritina Vázquez-Triñanes, Juan Carlos Lopez-Robledillo, Norberto Ortego-Centeno, Ana María Giménez-Arnau, Josep M. Campistol, Hafid Laayouni, Iñaki Ortiz de Landazuri, Jordi Yagüe, Eva Gonzalez-Roca, Anna Mensa-Vilaro, Oscar Fornas, Eduardo Ramos, Pablo Pelegrin, Ferran Casals, Juan I. Arostegui

## Abstract

**Objective:** *NLRP3* mosaicism is a well-established mechanism causing cryopyrin-associated periodic syndromes (CAPS). The number of reported patients with mosaicism is small, and the knowledge about the long-term disease behavior is limited. Herein we have assembled the largest cohort of individuals with *NLRP3* mosaicism reported to date to obtain additional evidence that strengthens the understanding of this disease.

**Methods:** Patients’ data were collected from their medical charts. Genetic analyses were performed using Sanger and next-generation sequencing. *In vitro* analyses determined the functional consequences of detected variants.

**Results:** Seventeen individuals with *NLRP3* mosaicism were enrolled, with 16/17 experiencing different CAPS phenotypes. An overrepresentation of late-onset forms was detected (37.5%). Overall, clinical manifestations, analytical results, and outcomes of treatments were markedly similar to those detected in patients with germline variants. A large mutational diversity was identified, with 16 different variants among 17 individuals. Two main patterns of mosaicism (extended vs myeloid-restricted) were detected, with the last one overrepresented in the late-onset group. The evaluation of mosaicism over time identified three different patterns, being the group with stable mosaicism the largest one.

**Conclusions:** Collected evidence supports the marked similarities among patients carrying somatic or germline *NLRP3* variants. The overrepresentation of *NLRP3* mosaicism in late-onset forms should be considered in patients with inflammatory manifestations starting in adulthood. Analysis of mosaicism at the biological level confirms the two known patterns of corporal distribution and reveals that mosaicism remains stable over time in most patients, but it may also vary during the course of the disease.

**Key Messages:** **What is already known about this subject?**

- Cryopyrin-associated periodic syndromes (CAPS) is a dominantly-inherited autoinflammatory disease with three different phenotypes of increasing severity across a continuous spectrum.
- The disease is a consequence of monoallelic, gain-of-function *NLRP3* variants leading to a constitutive hyperactivation of the NLRP3-inflammasome, with the subsequent IL-1β overproduction.
- Recent investigations have established the key role of *NLRP3* mosaicism in the pathogenesis of CAPS. However, the number of individuals with *NLRP3* mosaicism reported to date is small, and the data on the long-term behavior of the disease at clinical and biological levels are scarce.

**What does this study add?**

- We have assembled the largest cohort of individuals with *NLRP3* mosaicism reported to date to obtain novel evidence that strengthens the understanding of this syndrome.
- The clinical manifestations, the results of analytical tests, and the outcomes of treatments in patients with *NLRP3* mosaicism were in line with previous results collected from patients with germline *NLRP3* variants.
- We have detected a previously unidentified overrepresentation of late-onset forms among patients with *NLRP3* mosaicism.
- Genetic evidence supports for a large mutational diversity among individuals with *NLRP3* mosaicism. Despite the fact that, theoretically, any amino acid residue in the protein may be involved in mosaicism, the regions spanning the amino acid residues 300-310 and 560-570 concentrate the larger proportion of post-zygotic variants.
- Additional experiments confirmed the two main patterns of mosaicism distribution (myeloid-restricted vs extended) and that the degree of mosaicism remains stable in most patients but can also change over time.

**How might this impact on clinical practice?**

- Considering that genetic analyses represent the unique way to establish a CAPS diagnosis, the data presented here strongly suggest for an adequate analysis of the sequence reads of the *NLRP3* gene in candidate patients, which should include differential analyses for either germline and post-zygotic variants.
- The overrepresentation of late-onset forms among patients with *NLRP3* mosaicism should be taken into consideration by clinicians treating adult patients in the differential diagnosis of inflammatory diseases starting during adulthood.
- The evidence presented here support periodic evaluation of circulating *NLRP3* mosaicism to evaluate the disease behavior at biological level over time.

## Introduction

*NLRP3* encodes cryopyrin, a constitutive element of the large cytosolic complex called NLRP3-inflammasome (1). After proper activation, NLRP3-inflammasome activates caspase-1 that generates the active forms of different pro-inflammatory cytokines such as interleukin (IL)-1β (2). Gain-of-function (GoF) *NLRP3* variants cause the dominantly-inherited autoinflammatory disease (AID) known as cryopyrin-associated periodic syndromes (CAPS) (3). This disease encompasses three different phenotypes of increasing severity across a continuous spectrum. Familial cold autoinflammatory syndrome (FCAS) represents the mildest form, Muckle-Wells syndrome (MWS) exhibits intermediate severity, and chronic, infantile, neurologic, cutaneous and articular (CINCA) syndrome, also known as neonatal-onset multisystem inflammatory disease (NOMID), lies at the most severe end of the spectrum (3–4). CAPS-associated *NLRP3* variants lead to variable degrees of constitutive hyperactivation of the NLRP3-inflammasome (5–7). This hyperactive NLRP3-inflammasome leads to an IL-1β overproduction, which represents the rationale to treat CAPS with IL-1 inhibitors (8). Novel NLRP3 inhibitors based on the sulfonyl urea MCC950 compound also decrease inflammatory markers in CAPS, representing a future therapeutic approach for this condition (9).

From a genetic perspective, most CAPS-associated *NLRP3* variants are rare germline missense variants located in exon 4 (10). Some pathogenic variants lacking one or more of these features have been recently reported. In some instances, the variants were located outside exon 4, such as the p.Gly755Arg/Ala or p.Tyr859Cys/His variants, located in exon 5 and 7, respectively (10). In other cases, the *NLRP3* pathogenic variant was a post-zygotic, rather than germline, with a variant allele frequency (VAF) lower than 40%, leading to genetic mosaicism (11). CAPS patients due to *NLRP3* mosaicism remain a rarity with around 70 reported cases worldwide (11–24). Most of these patients have been described in case reports or exclusively from a genetic perspective, leading to a nearly complete absence of data on their long-term consequences, their responses to administered treatments, the functionality of their variant inflammasomes, or the longitudinal clonal dynamics of mosaicism.

The present work is a retrospective study collecting 17 unrelated individuals with variable degrees of *NLRP3* mosaicism. We aimed to generate new evidence about these patients by thoroughly describing their clinical and analytical features at diagnosis and during their follow-up, as well as the outcomes of administered treatments. Moreover, experimental studies were performed to characterize the variants’ pathogenicity, to investigate the mosaicism distribution and to assess the clonal dynamics of *NLRP3* mosaicism over time.

## Methods

### Study Participants

We retrospectively selected 17 individuals who carried an *NLRP3* mosaicism (VAF<44.1%) (25) among those received from Spanish medical centers, in 2005-2024, to undergo genetic analysis. Clinical data, results of analytical tests and outcomes of treatments were collected from the patient’s medical charts. Written informed consent for study enrollment was obtained from patients (>18 years) or patient’s parents (<18 years). The Ethical Review Board of Hospital Clínic approved the study (code HCB/2022/0855). All investigations were performed in accordance with the ethics standards of the 1964 Declaration of Helsinki and its later amendments. A complete description of the methods used for the different experimental studies and for statistical analyses is available in the Supplementary Appendix.

## Results

### Features of Enrolled Individuals

Seventeen individuals (7 males / 10 females) with post-zygotic *NLRP3* variants were enrolled. From them, 94.2% (16/17) were clinically symptomatic and diagnosed as suffering from different CAPS phenotypes: four CINCA-NOMID, six late-onset MWS (Lo-MWS), four MWS, one overlapping MWS-FCAS and one with undefined phenotype due to his young age at diagnosis (Supplementary Table S1). The remaining subject (P8) was an asymptomatic woman carrying a somatogonadal *NLRP3* mosaicism, who was reported elsewhere (26). At present, 14 individuals remained alive, whereas three had died as a consequence of neoplasia, a multiorgan failure secondary to sepsis, and a car crash, respectively.

Among patients, the mean age at disease onset was 26.2 years (range 0.1-76 years). Two extreme groups could be established on the basis of the age at disease onset: 6/16 patients (37.5%) in whom the disease started before the age of five years (early-onset group), and 6/16 patients (37.5%) in whom the disease manifested after the age of 45 years (late-onset group) (Figure 1A). The mean time of disease evolution was 27.1 years (range 2.0-72 years). The mean delay time from disease onset to genetic confirmation was 21.2 years (range 0.3-59 years), with a clear tendency for this delay to decrease among individuals born in more recent decades or among patients in the late-onset group (Figure 1B).

**Figure 1.**
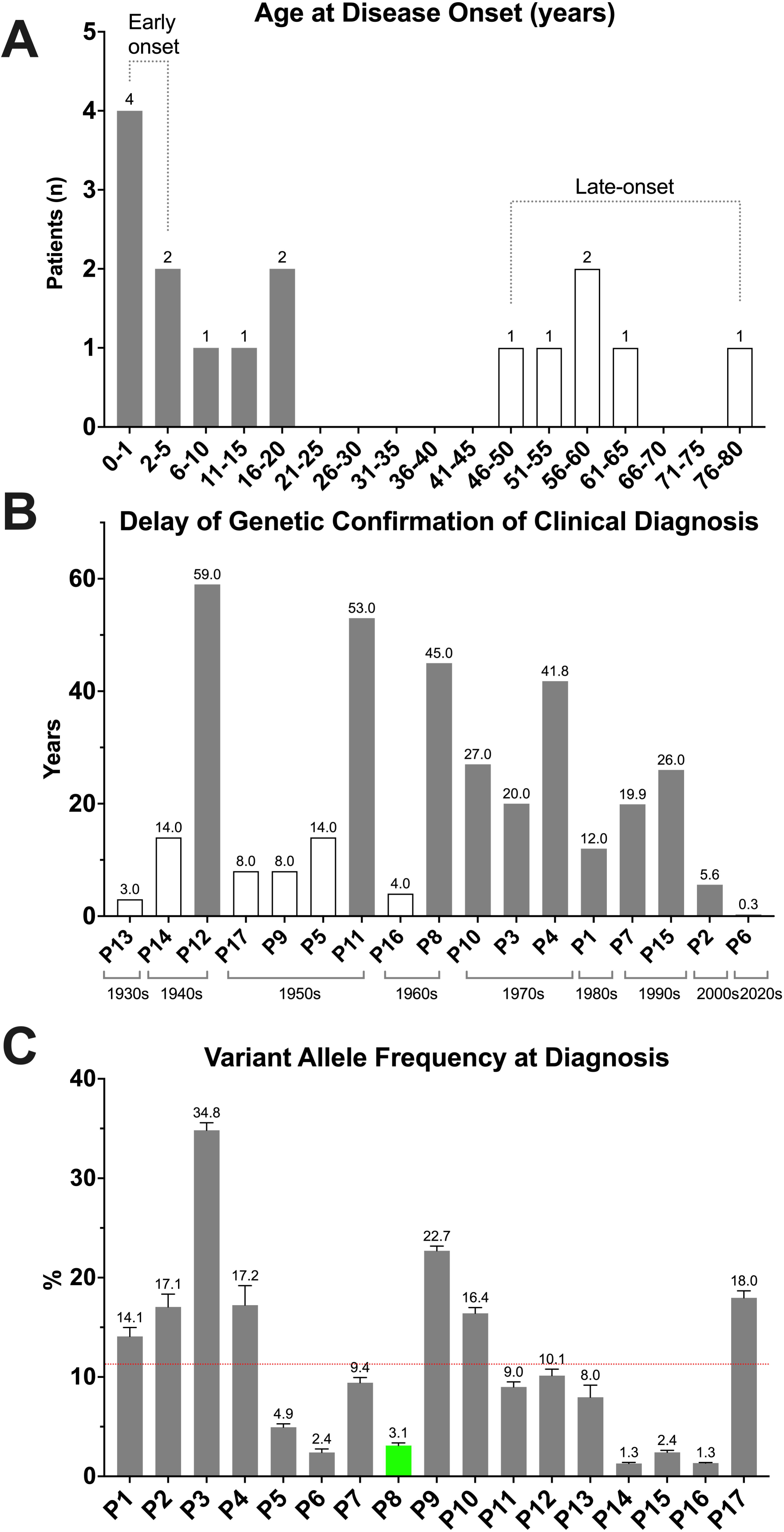
Panel A. Distribution of enrolled patients according to their age at disease onset. White bars indicate those patients belonging to the late-onset group. **Panel B**. Delay of diagnosis detected since disease onset until genetic confirmation. White bars indicate those patients belonging to the late-onset group. **Panel C**. Variant allele frequency (VAF) of detected *NLRP3* variants at diagnosis. Green bar indicates the asymptomatic individual, whereas gray bars represent the symptomatic patients. Horizontal dotted red line represents the mean VAF.

All patients displayed recurrent and simultaneously occurring inflammatory manifestations, frequently triggered by cold exposure (31%) or stress (31%). The most frequent manifestations at diagnosis included urticaria-like rash (100%), arthralgia (87%), recurrent fever (75%), arthritis (68%), and headache (56%) (Table 1). Before starting anti-IL-1 treatment, nearly all patients presented marked analytical perturbations. The most frequent hematological abnormalities included low mean hemoglobin concentration (14/16 patients; 87.5%), increased mean neutrophil count (14/16 patients; 87.5%), increased mean leukocyte count (12/16 patients; 75.0%), and increased mean platelet count (7/16 patients; 43.75%) (Supplementary Table S2, and Figures 2 and 3). Among biochemical tests, a moderate-to-high increase in erythrocyte sedimentation rate (ESR) and C-reactive protein (CRP) serum concentration was detected in nearly all patients (87.5% and 100%, respectively) (Supplementary Table S3 and Figure 4). As two different groups were identified based on age at disease onset, we performed a comparative analysis of their disease manifestations, with the results being shown in the Supplementary Table S4.

**Figure 2.**
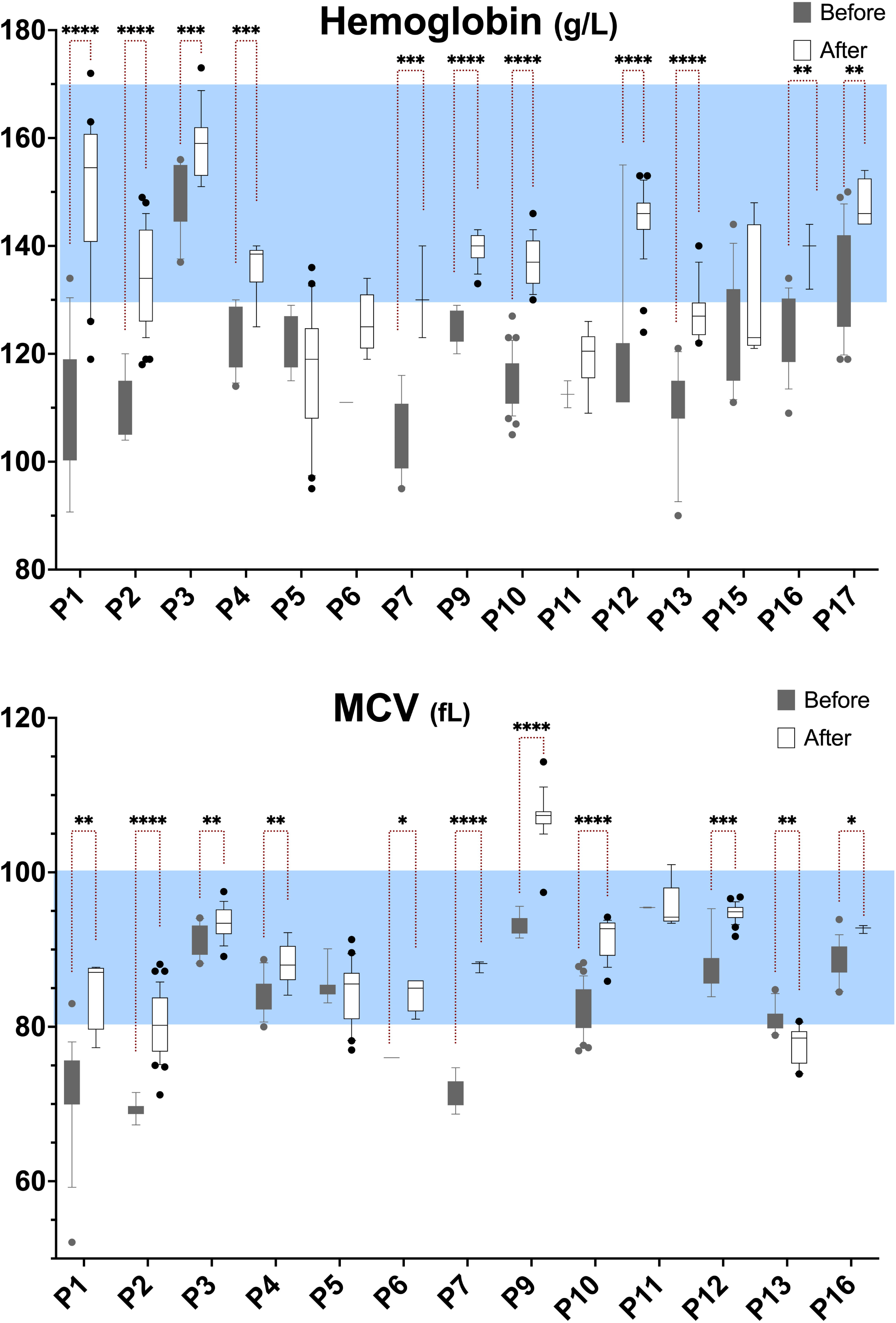
Results of hemoglobin concentration (Panel A) and mean corpuscular volume (Panel B) obtained before and during treatment with anti-IL-1 drugs. The blue rectangles indicate the normal reference range for each parameter. The boxplots show the 25^th^ to 75^th^ percentiles, the median line, and whiskers marking the 10^th^ to 90^th^ percentiles. *p<0.05, **p<0.01, ***p<0.001, ****p<0.0001.

**Figure 3.**
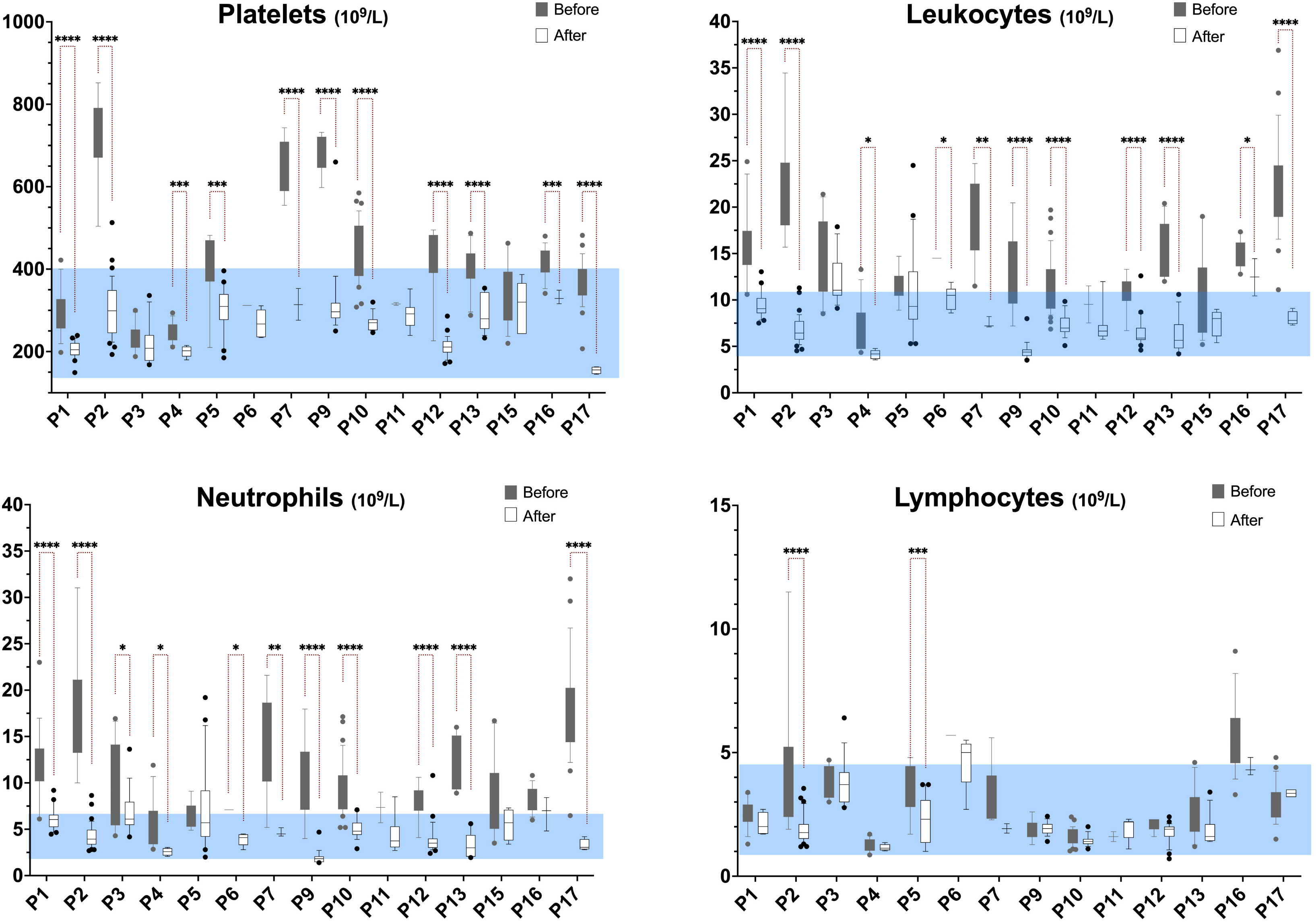
Results of platelet (Panel A), leukocyte (Panel B), neutrophil (Panel C) and lymphocyte (Panel D) counts obtained before and during treatment with anti-IL-1 drugs. The blue rectangles indicate the normal reference range for each parameter. The boxplots show the 25^th^ to 75^th^ percentiles, the median line, and whiskers marking the 10^th^ to 90^th^ percentiles. *p<0.05, **p<0.01, ***p<0.001, ****p<0.0001.

**Figure 4.**
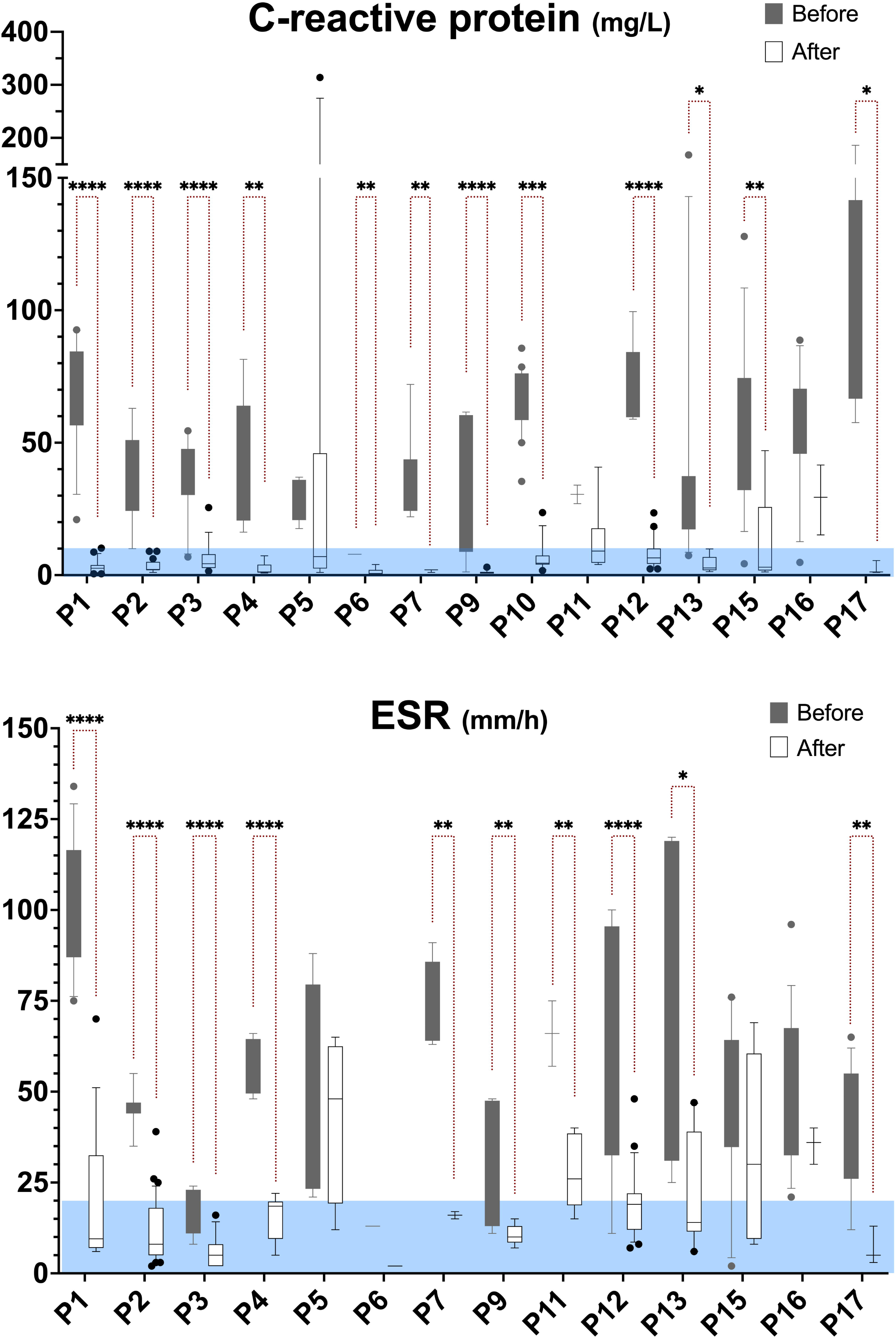
Results of serum C-reactive protein concentration (Panel A) and erythrocyte sedimentation rate (Panel B) obtained before and during treatment with anti-IL-1 drugs. The blue rectangles indicate the normal reference range for each parameter. The boxplots show the 25^th^ to 75^th^ percentiles, the median line, and whiskers marking the 10^th^ to 90^th^ percentiles. *p<0.05, **p<0.01, ***p<0.001, ****p<0.0001.

**Table 1.**
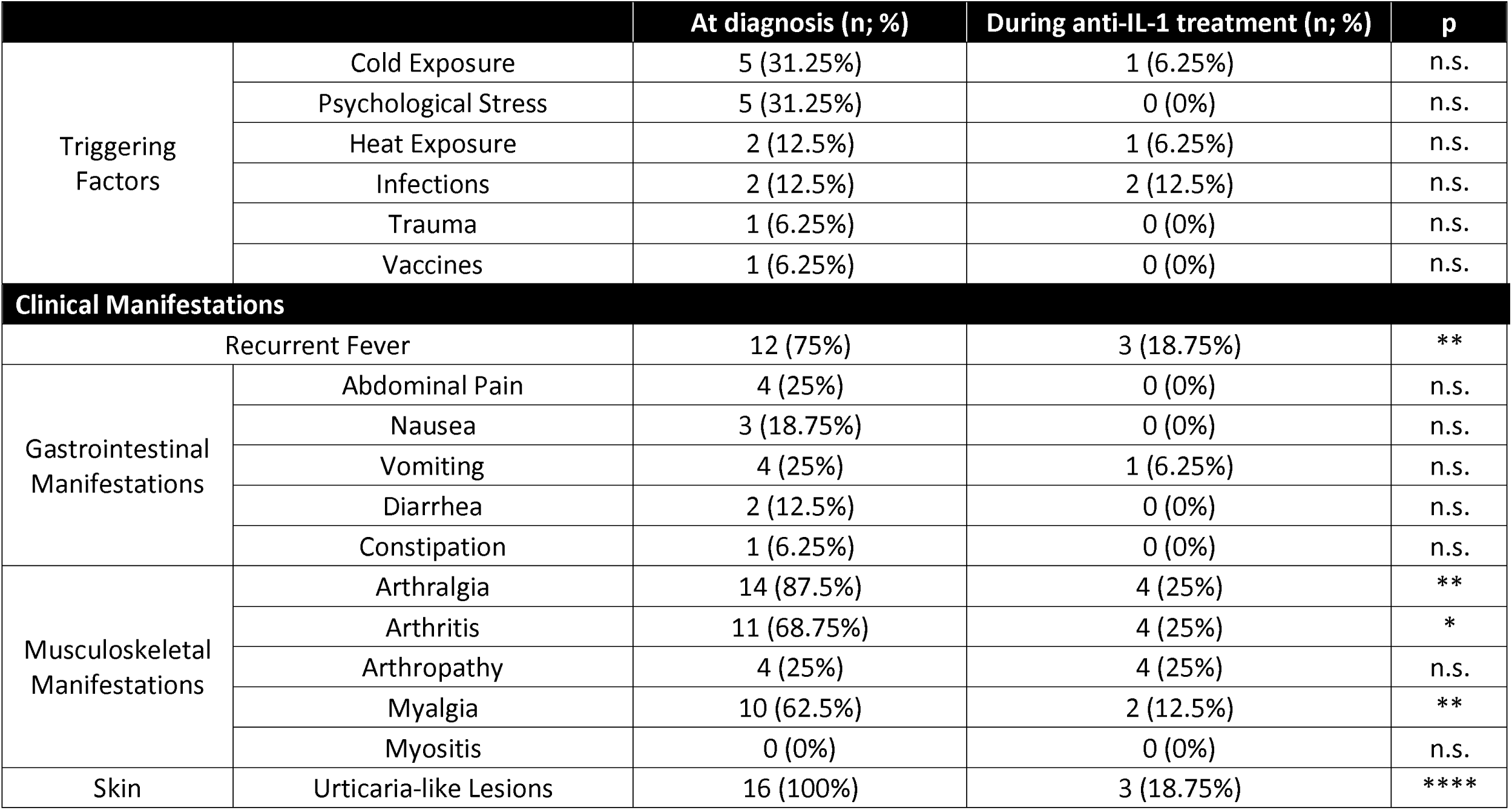
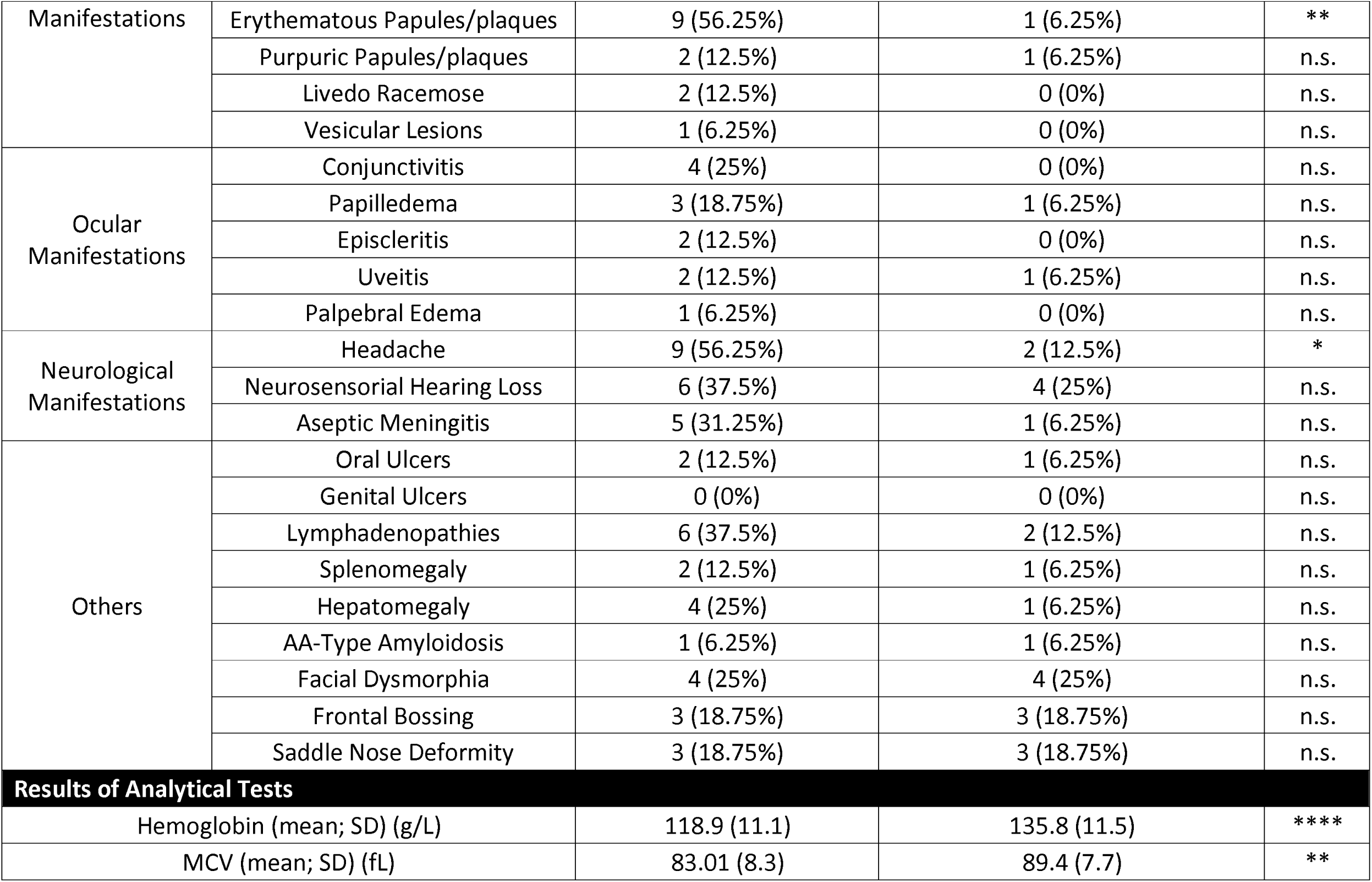
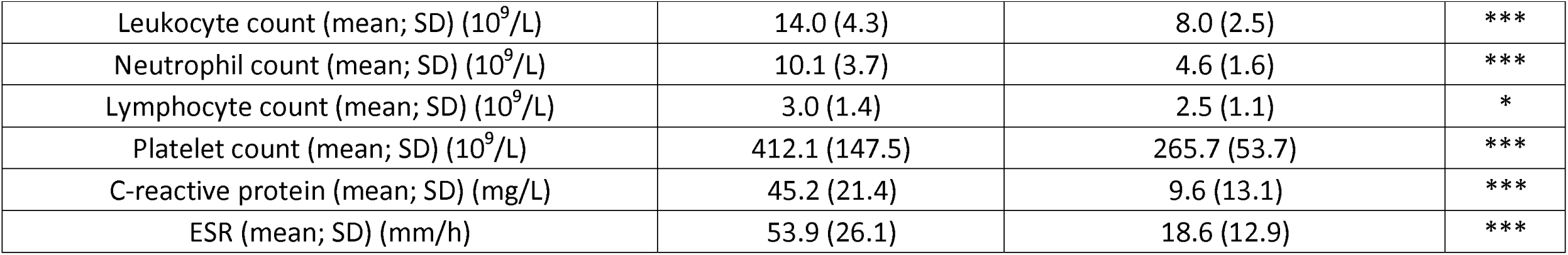
Clinical Features of Enrolled Patients at diagnosis and during treatment with anti-IL-1 drugs. Statistical analyses were performed using the McNemar test for the clinical characteristics and the paired t-student test for the results of analytical tests. *p<0.05, **p<0.01, ***p<0.001, ****p<0.0001. Abbreviations: MCV, mean corpuscular volume; ESR, erythrocyte sedimentation rate; n.s., not significant.

|  |  | At diagnosis (n; %) | During anti-IL-1 treatment (n; %) | p |
| --- | --- | --- | --- | --- |
| Triggering Factors | Cold Exposure | 5 (31.25%) | 1 (6.25%) | n.s. |
|  | Psychological Stress | 5 (31.25%) | 0 (0%) | n.s. |
|  | Heat Exposure | 2 (12.5%) | 1 (6.25%) | n.s. |
|  | Infections | 2 (12.5%) | 2 (12.5%) | n.s. |
|  | Trauma | 1 (6.25%) | 0 (0%) | n.s. |
|  | Vaccines | 1 (6.25%) | 0 (0%) | n.s. |
| **Clinical Manifestations** |  |  |  |  |
| Recurrent Fever |  | 12 (75%) | 3 (18.75%) | ** |
| Gastrointestinal Manifestations | Abdominal Pain | 4 (25%) | 0 (0%) | n.s. |
|  | Nausea | 3 (18.75%) | 0 (0%) | n.s. |
|  | Vomiting | 4 (25%) | 1 (6.25%) | n.s. |
|  | Diarrhea | 2 (12.5%) | 0 (0%) | n.s. |
|  | Constipation | 1 (6.25%) | 0 (0%) | n.s. |
| Musculoskeletal Manifestations | Arthralgia | 14 (87.5%) | 4 (25%) | ** |
|  | Arthritis | 11 (68.75%) | 4 (25%) | * |
|  | Arthropathy | 4 (25%) | 4 (25%) | n.s. |
|  | Myalgia | 10 (62.5%) | 2 (12.5%) | ** |
|  | Myositis | 0 (0%) | 0 (0%) | n.s. |
| Skin | Urticaria-like Lesions | 16 (100%) | 3 (18.75%) | **** |
| Manifestations | Erythematous Papules/plaques | 9 (56.25%) | 1 (6.25%) | ** |
|  | Purpuric Papules/plaques | 2 (12.5%) | 1 (6.25%) | n.s. |
|  | Livedo Racemose | 2 (12.5%) | 0 (0%) | n.s. |
|  | Vesicular Lesions | 1 (6.25%) | 0 (0%) | n.s. |
| Ocular Manifestations | Conjunctivitis | 4 (25%) | 0 (0%) | n.s. |
|  | Papilledema | 3 (18.75%) | 1 (6.25%) | n.s. |
|  | Episcleritis | 2 (12.5%) | 0 (0%) | n.s. |
|  | Uveitis | 2 (12.5%) | 1 (6.25%) | n.s. |
|  | Palpebral Edema | 1 (6.25%) | 0 (0%) | n.s. |
| Neurological Manifestations | Headache | 9 (56.25%) | 2 (12.5%) | * |
|  | Neurosensory Hearing Loss | 6 (37.5%) | 4 (25%) | n.s. |
|  | Aseptic Meningitis | 5 (31.25%) | 1 (6.25%) | n.s. |
| Others | Oral Ulcers | 2 (12.5%) | 1 (6.25%) | n.s. |
|  | Genital Ulcers | 0 (0%) | 0 (0%) | n.s. |
|  | Lymphadenopathies | 6 (37.5%) | 2 (12.5%) | n.s. |
|  | Splenomegaly | 2 (12.5%) | 1 (6.25%) | n.s. |
|  | Hepatomegaly | 4 (25%) | 1 (6.25%) | n.s. |
|  | AA-Type Amyloidosis | 1 (6.25%) | 1 (6.25%) | n.s. |
|  | Facial Dysmorphism | 4 (25%) | 4 (25%) | n.s. |
|  | Frontal Bossing | 3 (18.75%) | 3 (18.75%) | n.s. |
|  | Saddle Nose Deformity | 3 (18.75%) | 3 (18.75%) | n.s. |
| **Results of Analytical Tests** |  |  |  |  |
| Hemoglobin (mean; SD) (g/L) |  | 118.9 (11.1) | 135.8 (11.5) | **** |
| MCV (mean; SD) (fL) |  | 83.01 (8.3) | 89.4 (7.7) | ** |

|  |  |  |  |
| --- | --- | --- | --- |
| Leukocyte count (mean; SD) ($10^9/L$) | 14.0 (4.3) | 8.0 (2.5) | *** |
| Neutrophil count (mean; SD) ($10^9/L$) | 10.1 (3.7) | 4.6 (1.6) | *** |
| Lymphocyte count (mean; SD) ($10^9/L$) | 3.0 (1.4) | 2.5 (1.1) | * |
| Platelet count (mean; SD) ($10^9/L$) | 412.1 (147.5) | 265.7 (53.7) | *** |
| C-reactive protein (mean; SD) (mg/L) | 45.2 (21.4) | 9.6 (13.1) | *** |
| ESR (mean; SD) (mm/h) | 53.9 (26.1) | 18.6 (12.9) | *** |

### Outcomes of Medical Treatments

From disease onset until CAPS confirmation, all patients were treated with several anti-inflammatory drugs, including biological (median 4; range 1-8). The outcomes of these treatments were highly variable depending on the drug, and are detailed in Supplementary Table S5. Of note, only one patient (P9) received IL-1 inhibition empirically (anakinra; 100 mg q1d s.c.) before the genetic study on the basis of her clinical manifestations, which resulted in a positive response.

Once the genetic study confirmed the *NLRP3* mosaicism, all remaining patients started treatment with anti-IL-1 drugs. Anakinra was selected as the initial drug in 14/16 patients (87.5%) and canakinumab in 2/16 (12.5%). The response to anakinra was scored as effective in 13/14 patients (92.9%), and partial in one patient (P5). The positive responses included both the alleviation of most of the previous manifestations and the normalization of analytical perturbations (Table 1, Figures 2-4 and Supplementary Tables S2 and S3). Moreover, treatment with anakinra was well tolerated, with no serious adverse events detected, and with side effects at the site of injection as the most frequently observed adverse event. The intensity of these reactions was often mild-to-moderate, and drug discontinuation was only required in one patient (P14). During the follow-up, the initial treatment with anakinra was switched to canakinumab in six patients, due to partial response (n: 1), side effects at site injection (n: 1), and patient’s comfort (n: 4). All patients who received canakinumab also experienced a sustained alleviation of their clinical manifestations and normalization of analytical parameters, with no serious adverse events detected (Figures 2-4 and Supplementary Tables S2 and S3).

### Postzygotic *NLRP3* Variants

Genetic screening identified 16 different single nucleotide variants in the *NLRP3* gene. All these variants were located in the exon 4 and were predicted to lead to amino acid substitutions (Figure 5A and Supplementary Table S6). Among them, two variants were novel (p.Gly307Ala and p.Gly569Glu). Among the already reported variants, seven have been detected in both germline and post-zygotic states, while the remaining eight have been exclusively reported as post-zygotic variants. According to the ACMG/AMP guidelines for variant classification, all variants were classified as pathogenic/likely pathogenic (Supplementary Table S6). At the time of the first genetic evaluation, the mean VAF was 11.3% (range 1.3-34.8%), being the frequency of mutant allele in each individual compatible with that expected for post-zygotic variants causing mosaicism (Figure 1C) (25). Additional genetic analyses performed on the healthy parents of four patients (P1, P2, P6 and P7) were all negative (data not shown), thus confirming the *de novo* nature of the variants in these families.

**Figure 5.**
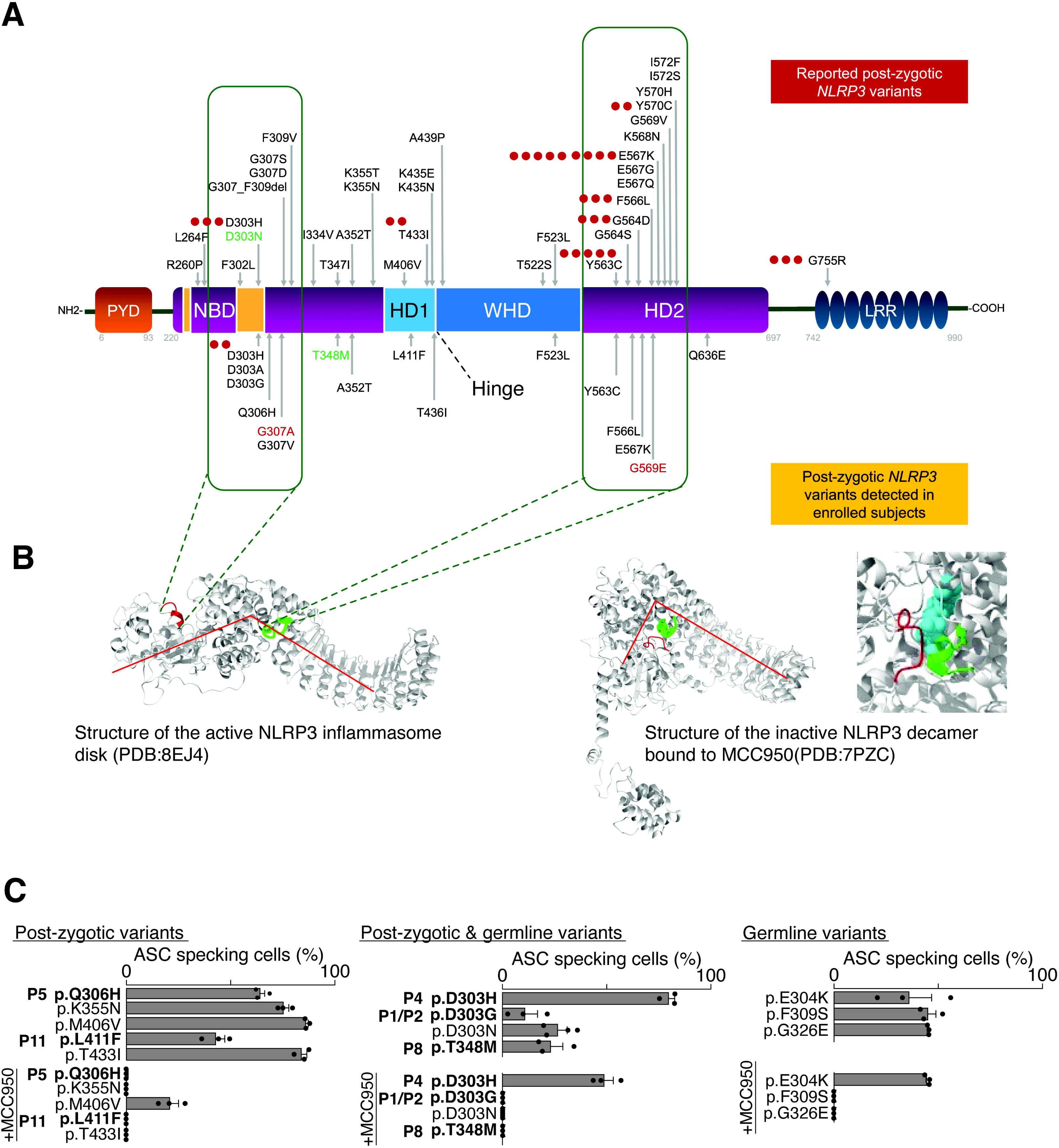
Domain organization and functionality of cryopyrin and localization of known post-zygotic *NLRP3* pathogenic variants. **Panel A.** Above the protein scheme are indicated those post-zygotic *NLRP3* variants reported elsewhere, and below the protein scheme those post-zygotic variants detected in enrolled individuals. The novel *NLRP3* variants are highlighted in red letters, whereas those post-zygotic variants detected in healthy individuals are highlighted in green letters. Boxes indicates the regions spanning the amino acid resides 300-310 (left) and 560-570 (right), where the post-zygotic *NLRP3* variants concentrate. Red dots indicate the number of patients identified as carriers of the concrete post-zygotic *NLRP3* variant. PYD: Pyrin domain; NBD: Nucleotide binding domain (Walker A and B marked in orange); HD: Helix domain; WHD: Wind helix domain; LRR: Leucine rich repeat domain. **Panel B.** Chain A of the NLRP3 cryo-EM structure, corresponding to the active inflammasome disk (PDB: 8EJ4, left) and the inactive decamer bound to MCC950 (PDB: 7PZC, right), regions corresponding to the regions where the post-zygotic *NLRP3* variants are concentrated are denoted in red and green. **Panel C.** Percentage of ASC-specking HEK293T cells expressing different *NLRP3* variants as indicated in the presence or absence of the NLRP3 inhibitor MCC950.

### *In vitro* Functional Characterization of NLRP3 Variants

Two regions inside the NACHT domain of NLRP3 concentrate most of the post-zygotic variants. These regions were located between the amino acid residues 300-310, spanning the Walker B motif for nucleotide hydrolysis, and 560-570, inside the HD2 domain (Figure 5A). In the inactive NLRP3 decamer structure, these regions appear adjacent and interacting, close to the MCC950 binding pocket. Therefore, some amino acid change in these regions will potentially destabilize the inactive structure (Figure 5B). Furthermore, in the active NLRP3 structure of the inflammasome disk, these two regions fall apart (Figure 5B). To characterize the functional consequences of detected variants, we analyzed their ability to form active inflammasomes (Figure 5C). We found that the five analyzed *NLRP3* variants presented a differential percentage of inflammasome activation, ranging from 10 to 70% of ASC-specking cells (Figure 5C), in line with previous results for germline variants (6). Of note, we were unable to correlate *in vitro* activity of the different *NLRP3* variants with the CAPS phenotype, even for those variants only found in post-zygotic status. The specific NLRP3 inhibitor MCC950 resulted in a complete abolishment of ASC-specking cells when the majority of post-zygotic *NLRP3* variants were expressed, except for the p.D303H that presented only a partial inhibitory effect (Figure 5C). These results suggest that CAPS patients with the post-zygotic *NLRP3* variants would be candidates to be treated with sulfonyl urea-derivatives drugs, albeit specific dosage will require further studies.

### Longitudinal Clonal Dynamics of Mosaicism

To investigate the stability of mosaicism over time, we collected blood samples at different time points from 16 individuals (15 patients and the asymptomatic individual). The number of collected samples per individual ranges between two and four, whereas the time interval from the first to the last collected sample was highly variable (range 2-259 months). We performed the formal analysis of mosaicism stability in 13 individuals, excluding those patients in whom the time interval between the first and last collected sample was shorter than 12 months (P5, P15 and P17). The results of these experiments indicate that *NLRP3* mosaicism remained stable in seven individuals (7/13; 53.8%), increased in three individuals (3/13; 23.1%), and decreased in three individuals (3/13; 23.1%) (Figure 6 and Supplementary Table S7). The higher increase in VAF was detected in P10 (increase from 16.4% up to 33.0% in 255 months; Supplementary Figure S1) and in P13 (increase from 8% up to 13.5% in 104 months; Supplementary Figure S2). By contrast, the largest decreases in VAF were detected in P9 (decrease from 22.7% up to 2.2% in 88 months; Supplementary Figure S3), P1 (decrease from 14.1% up to 7.9% in 143 months; Supplementary Figure S4) and P2 (decrease from 17.1% up to 10.0% in 209 months; Supplementary Figure S5).

**Figure 6.**
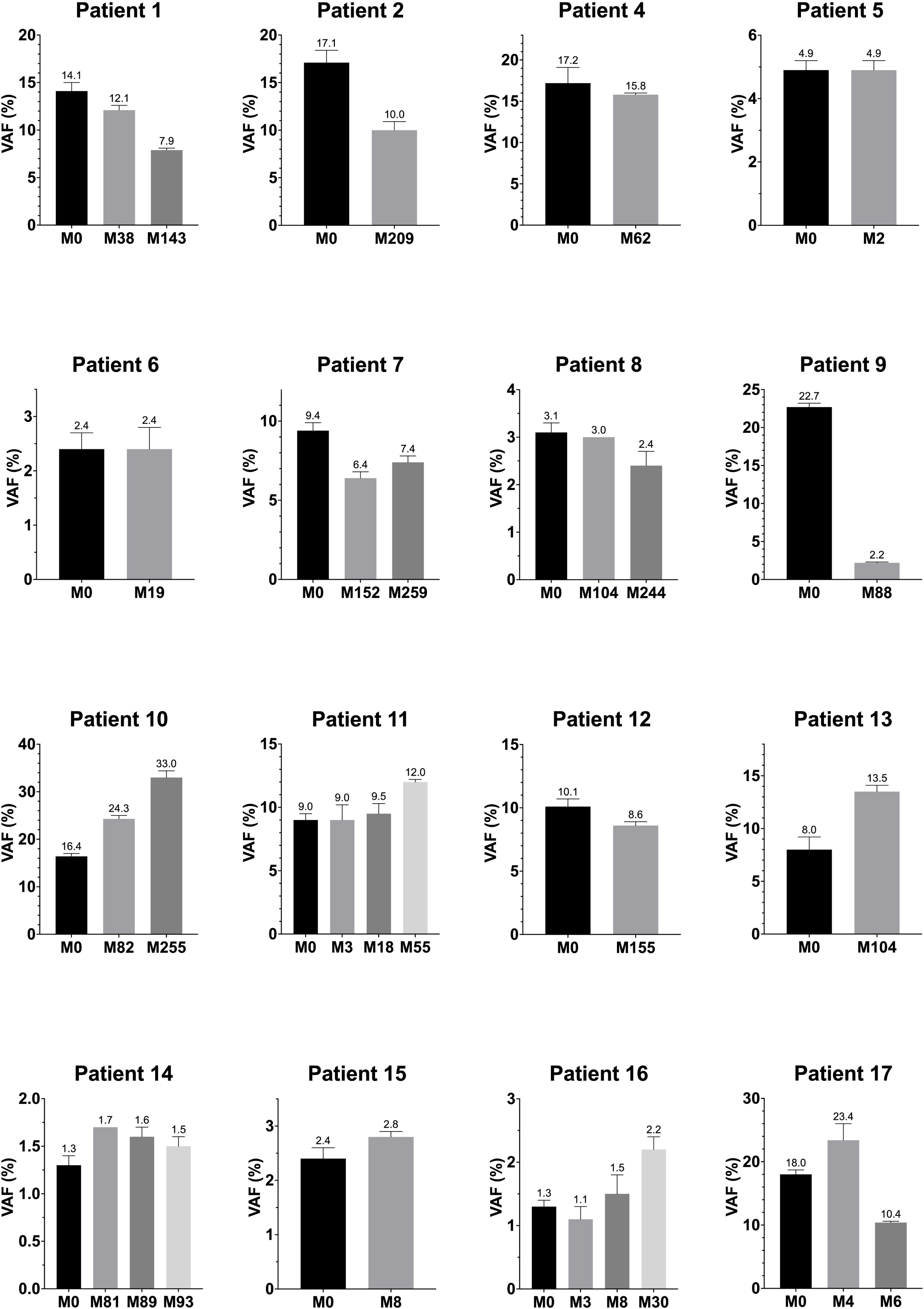
Clonal dynamics of *NLRP3* mosaicism over time. Every panel represents the evolution of the frequency of the mutant *NLRP3* allele in each analyzed individual over time. X-axes represent the time in months (M) from diagnosis to sample collection, with M0 being the results obtained at genetic diagnosis. Abbreviations: VAF, variant allele frequency.

### Distribution of *NLRP3* Mosaicism

To draw conclusions about the timing of the post-zygotic mutational event, we investigated the mosaicism distribution across different tissues. We first evaluated the presence of the *NLRP3* variants in different leukocyte subpopulations. For this purpose, blood samples from 14 subjects were collected and their leukocyte subpopulations were isolated. Subsequent amplicon-based deep sequencing (ADS) analyses using genomic DNA from each isolated subpopulation revealed two patterns of mosaicism. On one side, the main group (9/14; 64.3%) included those individuals in whom the *NLRP3* variant was detected in both myeloid and lymphoid cells at similar percentages (Figure 7 and Supplementary Table S8). The second group (5/14; 35.7%) included individuals (P5, P9, P13, P16, P17) in whom the *NLRP3* variant was primarily detected in myeloid cells, with little to no involvement of lymphoid cells or extra-hematopoietic tissues (Supplementary Table S8). These results might be compatible with a myeloid-restricted *NLRP3* mosaicism in line with previous reports (16, 17, 19).

**Figure 7.**
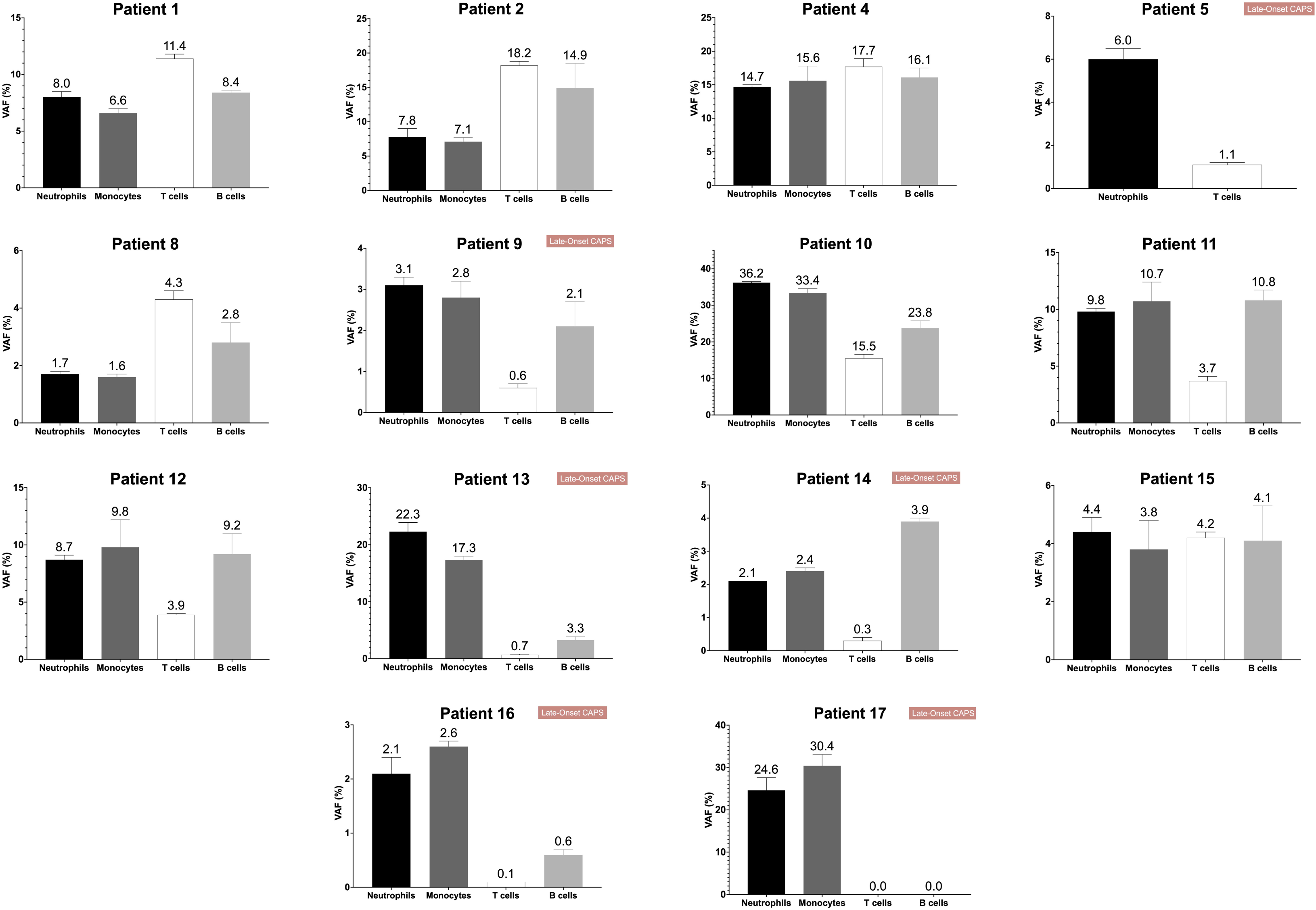
Patterns of Mosaicism Distribution in Leukocyte Subpopulations. Each panel shows the results of amplicon-based deep sequencing using DNA extracted from isolated leukocyte subpopulations. Each bar represents the mean variant allele frequency (VAF) from three independent experiments in the indicated leukocyte subpopulation and the whiskers on top indicate the standard deviation (SD).

## Discussion

Gene mosaicism is a biological phenomenon largely investigated in malignant diseases, where normal and neoplastic cells can be isolated and individually analyzed (27). The relevance of mosaicism in the pathogenesis of inherited diseases started to be elucidated when NGS-based methods arrived into the clinics, based on their ability to detect mutant alleles at low frequencies (28). Among rheumatological diseases, the key role that mosaicism plays in the pathogenesis of certain AID is currently well established, with VEXAS and CAPS as the prototypical examples (29, 30).

Around 70 patients with CAPS due to *NLRP3* mosaicism have been described to date (11–24). Most of them were described in case reports, with no longitudinal evaluation of clinical or biological parameters, while others were merely described from a genetic point of view. Herein, we have assembled the largest cohort to date of individuals with *NLRP3* mosaicism, aiming to fill potential gaps in the existing knowledge on this topic. From a clinical perspective, the large proportion of patients with late-onset disease (35.3%) in our cohort should be highlighted, as opposed to the classical conception that CAPS always start during pediatric ages (3). This particular information should be taken into consideration by clinicians treating adult patients in the differential diagnosis of suspected AID starting during adulthood. Overall, the manifestations and analytical results among patients carrying *NLRP3* mosaicism do not differ substantially from those reported in patients carrying germline mutations (3, 23). However, when analyzing the early-onset and late-onset groups of patients with *NLRP3* mosaicism, we found some particular differences beyond the age of disease onset. Thus, lymphadenopathies and some structural manifestations (arthropathy, dysmorphic features) seem to concentrate in the early-onset group. By contrast, myalgias or stress as a trigger are more frequently detected among patients in the late-onset group, with these results consistent with previously reported data (19).

The outcomes of treatments administered before confirming the CAPS diagnosis were characterized by the absence of a clear response to most of them, with the unique exception of oral corticosteroids. However, once CAPS diagnosis was established, all patients started IL-1 blockade, with patterns of response and safety nearly identical to those previously described in patients with germline *NLRP3* variants (8, 23, 31–33). Of note, *in vitro* functional analysis here performed found that the analyzed post-zygotic *NLRP3* variants are susceptible to be inhibited with novel NLRP3 inhibitors of the sulfonyl urea compounds, aligning with the data obtained in a Phase 2 clinical trial that enrolled patients with germline *NLRP3* variants (9).

From a genetic perspective, evidence already published, along with the data presented here, clearly supports the existence of significant genetic heterogeneity among CAPS patients with mosaicism as 45 different post-zygotic *NLRP3* variants have been identified in a total of 74 patients (Figure 5A). Moreover, different available evidence supports a GoF behavior of the *NLRP3* variants detected in our patients, including the positive response to anti-IL-1 drugs and *in vitro* functional studies that showed that some of these variants activate the NLRP3-inflammasome in a similar manner to other well-known germline pathogenic variants. This genetic diversity of post-zygotic *NLRP3* variants is similar to previous results obtained in the NLRC4-associated AID, where four different post-zygotic *NLRC4* variants were detected among the five reported patients (34–37). In contrast, these results are markedly different to the genetic homogeneity detected in patients with Blau syndrome, where all patients with mosaicism carry the p.Arg334Gln/Trp *NOD2* variants (25, 38, 39), or with VEXAS, where 85-90% of the patients carry the p.Met41Leu/Val/Thr *UBA1* variants (30, 40–42). We obtained additional interesting data about the detected *NLRP3* variants. With regard to their location, the results indicate that any amino acid residue may be theoretically involved in mosaicism, but two regions concentrate a larger proportion of post-zygotic variants. The first region spans the amino acid residues 300-310, a well-known functional domain of the protein that includes the Walker B motif, which concentrates 26.7% (12/45) of post-zygotic *NLRP3* variants known to date (Figure 5A). The second region spans the amino acid residues 560-570, which concentrates 31.1% (14/45) of known post-zygotic *NLRP3* variants. Of note, this region accumulates most of the variants detected in the late-onset forms, including those enrolled in this work (3/6; 50%) and those previously reported (19). Moreover, 64.4% (29/45) of known post-zygotic *NLRP3* variants have been identified exclusively as mosaics, with no germline counterpart. This observation leads to the hypothesis that these variants, in germline status, may provoke a severe intrauterine activation of the NLRP3-inflammasome leading to embryonic or fetal death. This hypothesis is supported by animal CAPS model with germline *NLRP3* variants, which present elevated embryonic lethality (43, 44), and by previous evidence reported in certain inherited diseases such as Proteus syndrome (45) and incontinentia pigmenti (46).

The longitudinal clonal dynamics of *NLRP3* mosaicism have been scarcely addressed in previous reports, with the main evidence coming from the British and the Italian cohorts (18, 19). The British cohort included exclusively patients with late-onset CAPS, and the authors found that the degree of *NLRP3* mosaicism in peripheral blood remained stable in most of the patients during a median of follow-up of three years. The unique exception was a female patient in whom an increase in VAF from 5.1% at diagnosis to 45% twelve-years later was detected in the absence of a proliferative disease (19). The Italian group analyzed exclusively patients with the early-onset form and they found that the *NLRP3* mosaicism remained stable in 75% (3/4) of the patients, whereas it decreased in the remaining one (18). This evidence suggested that *NLRP3* mosaicism may vary over time, but it came from a limited number of patients. In order to provide further data on this topic, we evaluated the clonal dynamics of *NLRP3* mosaicism in 16 individuals, including early- and late-onset forms and an asymptomatic individual. The analysis was performed on 13 patients for whom more than 12 months elapsed between the first and last collected blood samples. The results revealed three patterns of mosaicism over time. The main group (53.8%) included those individuals in whom the mosaicism remained stable, in line with previous data (18, 19). A second pattern included those patients in whom the degree of mosaicism increased in absence of a proliferative disease and despite a good response to anti-IL-1 drugs. These patients mirror the aforementioned British patient, with the underlying reasons for the expansion of the mutant clone yet to be elucidated. The final group included those patients in whom a marked decrease of the circulating mosaicism was detected. As patients belonging to the three patterns experienced similar successful responses to anti-IL-1 drugs, this treatment could not explain by itself the decrease of mosaicism detected in the last group. An attractive hypothesis might be the progressive exhaustion of the mutant clone over time. Theoretically, in the case that the mutant clone disappears completely, this might lead to the spontaneous curation of the disease, a phenomenon still undescribed in CAPS. In VEXAS syndrome, different groups have reported a similar phenomenon of mosaicism decrease over time as a consequence of a specific treatment. These groups reported marked decreases of the frequency of *UBA1* mutant allele when patients were successfully treated with hypomethylating agents, but no changes when the same patients were treated with anti-inflammatory drugs including IL-1-, IL-6- or TNF-inhibitors (42, 47, 48). Interestingly, the pathogenic *UBA1* mutations in VEXAS are restricted to myeloid cells, which also represent the relevant cell types in CAPS pathogenesis (16, 17). Although hypomethylating drugs are not formally indicated for the treatment of CAPS, an open question remains as to whether these drugs could lead to successful clinical responses and/or a reduction in circulating *NLRP3* mosaicism in CAPS, as has been reported in VEXAS. Overall, the evidence collected here confirms the previous observation of potential variation of mosaicism over time, strongly supporting its periodic evaluation in each patient to assess the disease evolution at a biological level.

The distribution of mosaicism in leukocyte subpopulations has been previously reported in occasional patients (12, 13, 16–19). Herein, we analyzed this issue in 14 patients, representing the largest cohort to date for these studies. The results confirmed the existence of two different patterns. The main group (9/14; 64.3%) includes those patients in whom the *NLRP3* variant was detected at similar levels in myeloid and lymphoid cells, being highly probable that the hematopoietic stem cells are also carriers of the variant. This observation strongly suggests an early occurrence of the mutational event leading to mosaicism in these patients, a conclusion that is also supported by the early-onset of the disease detected among these patients and by the presence of the *NLRP3* variant in tissues derived from other embryonic layers as revealed by analyses of nail tissues. The second pattern of mosaicism is a minority (5/14; 35.7%) and includes those patients in whom the *NLRP3* variant was primarily or entirely restricted to myeloid cells. This aligns with the fact that monocytes are the main cell type inducing constitutive activation of NLRP3-inflammasomes in CAPS (49). Interestingly, this pattern of mosaicism is overrepresented in patients with late-onset CAPS, and strongly suggests that the mutational event occurred during adulthood. Of note, this myeloid-restricted pattern of mosaicism has also been described in VEXAS syndrome, which represents the prototypical late-onset AID (29, 30, 42). Despite the different time when the mutational event occurred, no differences on the clinical manifestations, analytical results or outcomes of treatments have been detected between the two groups beyond the age at disease onset.

An interesting issue about *NLRP3* mosaicism is its potential presence in asymptomatic individuals. The present work includes a healthy woman with a long-lasting somatogonadal *NLRP3* mosaicism who was the subject of a previous report (26). Another similar case, a male carrying the p.Asp303Asn *NLRP3* variant as somatogonadal mosaicism, was also reported by our group (25). From a clinical perspective, the identification of this type of mosaicism has two different consequences. On one side, in the genetic counseling process of families in which a *de novo* variant has been detected, it is currently mandatory to analyze the potential presence of mosaicism in the asymptomatic parents by using methods with high sensitivity (i.e. ADS or digital droplet PCR) to correctly assess the risk of transmitting the mutant allele to the offspring in a future pregnancy. On the other side, those healthy individuals with mosaicism should be periodically evaluated from clinical and genetic points of view. Despite previous data and the results shown here pointing at the degree of mosaicism remaining stable over time as the most probable scenario, which would result in the subject remaining asymptomatic, there is also the possibility that the mutant clone may expand as the subject ages by either myeloproliferative disorders or by the phenomenon known as clonal hematopoiesis of indeterminate potential (50). If this happens, there is the possibility that the subject starts developing clinical symptoms similar to those detected among patients in the late-onset group, triggering the need to start treatment with anti-IL-1 drugs.

The main strengths of our study are the large number of individuals with *NLRP3* mosaicism coming from a single country enrolled for both clinical evaluation and experimental investigations, as well as the extensive collection of samples over a prolonged period to assess the clonal dynamics of mosaicism over time. The main limitations of our study stem from its retrospective nature, which made it difficult to obtain some of the oldest data due to their unavailability in electronic health records. Additionally, the loss of follow-up for some patients between their diagnosis and manuscript preparation, prevented the collection of further data.

In summary, we have built the largest cohort of patients with CAPS due to *NLRP3* mosaicism to describe in detail their clinical features, the outcomes of administered treatments and to investigate the clonal dynamics of mosaicism over time and its corporal distribution. The patients’ clinical manifestations, the results of different laboratory tests, and the outcomes of administered treatments were in line with previous reports, with the only exception of the large number of enrolled patients presenting with late-onset disease. From a genetic perspective, we confirm the mutational diversity among enrolled individuals and describe the main patterns of mosaicism distribution (myeloid-restricted vs extended). We also confirm that the degree of mosaicism remains stable in most patients but can also change over time, which may have clinical implications, with an increase leading to disease worsening, and a decrease possibly resulting in improvement or even spontaneous resolution.

## Data Availability

**Data sharing statement**
The data included in the present work are, by definition, deidentified, and are available upon reasonable request to the corresponding author. Data shall only be made available after approval by the study contributors of a submitted research proposal, with investigator support and after a signed data access agreement.

## Acknowledgments

The authors thank the patients and their families for their collaboration in this study and Nerea Moreno-Ruiz PhD for her assistance with manuscript revision.

## Contributorship

- Study Conception and Design: NB, PP, FC and JIA.
- Methodology-Data collection: NB, JMM, JLC-R, DC, AS, OL, NP-F, EB, SJ-T, AR, MAn, JR-M, MAp, DP, MTS-C, JML-R, MC-P, LIG-G, CP, JMG-RdeM, EGdelaF, AB, EI, JG-R, CV-T, JCL-R, NO-C, AMG-A, JMC, ER, JIA.
- Methodology-Samples Collection: NB, JMM, JLC-R, DC, NP-F, SJ-T, MA, LF-D, JR-M, MAp, JG-H, DP, LIG-G, JMG-RdeM, EGdelaF, EI, CV-T, AMG-A, JMC, ER, JIA.
- Methodology-Genetic Studies: NB, VF, MCA, SP, IOdeL, JY, EG-R, AM-V, FC, JIA
- Methodology-Leukocyte subpopulation isolation: NB, VB, SP, MD, AV, DL, OF, JIA
- Methodology-In vitro NLRP3-inflammasome experiments: LH-N, DA-B, JG-V, PP
- Methodology-Statistical Analyses: HL
- Formal Analysis: All authors
- Writing (original draft): NB and JIA
- Writing (revising, review, and editing): All authors
- All authors read and approved the final manuscript. JIA acts as guarantor for the study, had access to all data and controlled the decision to publish.

## Data sharing statement

The data included in the present work are, by definition, deidentified, and are available upon reasonable request to the corresponding author. Data shall only be made available after approval by the study contributors of a submitted research proposal, with investigator support and after a signed data access agreement.

## Patient and Public Involvement

Patients and/or the public were not involved in the design, conduct, reporting or dissemination plans of this research.

