## Supplementary Appendix for "Clinical Features, Outcomes of Treatments, Inflammasome Function and Longitudinal Clonal Dynamics into *NLRP3* Mosaicism: Evidence from the Largest Cryopyrin-associated Periodic Syndromes Cohort to Date"

Nuria Bonet, Jose M. Mascaro Jr, Laura Hurtado-Navarro, Diego Angosto-Bazarra, , Jose Luis Callejas-Rubio, Daniel Clemente, Alejandro Souto, Olalla Lima, Natalia Palmou-Fontana, Eulalia Baselga, Santiago Jiménez-Treviño, Agustin Remesal, Marta Andreu-Barasoain, Luis Fernandez-Dominguez, Josep Riera-Monroig, Maria Aparicio, Juan Garcia-Herrero, David Pesqué, Maria Teresa Sanchez-Calvin, Jose Miguel Lezana-Rosales, Maria Correyero-Plaza, Julio Garcia-Villalba, Victor Bolaño, Sara Peiro, Mar Diaz, Alexandru Vlagea, Daniel Lorca, Virginia Fabregat, Maria Carmen Anton, Susana Plaza, Luis Ignacio Gonzalez-Granado, Concepción Postigo, Jose Maria Garcia-Ruiz de Morales, Enrique Gómez de la Fuente, Estibaliz Iglesias, Javier Gomez-Roman, Caritina Vázquez-Triñanes, Juan Carlos Lopez-Robledillo, Norberto Ortego-Centeno, Ana María Giménez-Arnau, Josep M. Campistol, Hafid Laayouni, Iñaki Ortiz de Landazuri, Jordi Yagüe MD, Eva Gonzalez-Roca, Anna Mensa-Vilaro, Oscar Fornas, Eduardo Ramos, Pablo Pelegrin, Ferran Casals, Juan I. Arostegui

**Table of Contents**

**Supplementary Methods**

**Supplementary Tables and Legends**

- Supplementary Table S1. Main Clinical Manifestations of Enrolled Patients.
- Supplementary Table S2. Results of Hematological Parameters of Enrolled Patients Before and During Treatment with anti-IL-1 Drugs.
- Supplementary Table S3. Results of Biochemical Parameters of Enrolled Patients Before and During Treatment with anti-IL-1 Drugs.
- Supplementary Table S4. Clinical Features of Patients with Early-onset CAPS versus Late-onset CAPS.
- Supplementary Table S5. Outcome of Treatments in Enrolled Patients.
- Supplementary Table S6. Classification of Post-zygotic *NLRP3* Variants.
- Supplementary Table S7. Clonal Dynamics of *NLRP3* Mosaicism Over Time.
- Supplementary Table S8. Distribution of Mosaicism.
- Supplementary Table S9. List of analyzed genes associated with autoinflammatory diseases.

**Legends of Supplementary Figures**

- Supplementary Figure S1. Sanger chromatograms showing clonal dynamics of NLRP3 mosaicism in P10.
- Supplementary Figure S2. Sanger chromatograms showing clonal dynamics of NLRP3 mosaicism in P13.
- Supplementary Figure S3. Sanger chromatograms showing clonal dynamics of NLRP3 mosaicism in P9.
- Supplementary Figure S4. Sanger chromatograms showing clonal dynamics of NLRP3 mosaicism in P1.
- Supplementary Figure S5. Sanger chromatograms showing clonal dynamics of NLRP3 mosaicism in P2.

**References of Supplementary Appendix**

**Supplementary Methods**

**DNA extraction, quantification, and quality control**

Blood samples were collected in EDTA- and heparin-containing tubes. Nail clippings were obtained after thorough hand cleaning to prevent potential contamination by skin scratching. Genomic DNA of samples collected from study participants was extracted using the QIAmp DNA Blood Mini Kit or the QIAamp DNA Investigator Kit (both from QIAgen, Germany) according to the manufacturer’s instructions. DNA concentration and quality were assessed using NanoDrop (Thermo Fisher Scientific, USA) and Qubit (Life Technologies, USA) fluorometers. A260/A280 ratios of 1.8 to 2.0 and A260/A230 ratios of >1.5 were accepted.

**Targeted gene panel sequencing**

AID-associate genes (Supplementary Table S9) were analyzed using a targeted gene panel (TGP) sequencing approach. Briefly, the TGP library was generated in an Access Array System 48.48 platform (Fluidigm, USA). Library preparation and quality control were performed according to manufacturers’ instructions, and sequencing was performed on a NextSeq platform (Illumina, USA). Reads were mapped against reference genome GRCh38 using the BWA software (1), and subsequent analyses were performed using the SeqNext software (JSI Medical Systems, Germany). Detected variants were classified according to the consensus recommendations of the American College of Medical Genetics and Genomics (ACMG) and the Association for Molecular Pathology (AMP) (2).

**Amplicon-based deep sequencing**

The VAF of post-zygotic *NLRP3* variants was determined using the Amplicon-based deep sequencing (ADS) method as previously described (3). Briefly, amplicons covering the target regions of the *NLRP3* gene (RefSeq: NM_001243133.2) were generated by in-house designed PCR. Deep sequencing was performed on an S5XL platform (IonTorrent, Life Technologies, USA), with coverage >1000x. Reads were mapped against the GRCh38 using BWA, and variants were subsequently analyzed using the Integrative Genomics Viewer (4). The VAF for each experiment was calculated as the proportion of variant reads to total reads and expressed as a percentage.

**Isolation of human leukocyte subpopulations**

The main leukocyte subpopulations were isolated using either a flow cytometry approach (before the COVID19 pandemic) or an immunomagnetic method. Fresh whole blood samples were collected in EDTA- and heparin-containing tubes, and peripheral blood mononuclear cells (PBMCs) were isolated via Ficoll-Paque Premium (Cytiva, Freiburg, Germany). For cell sorting, whole blood or PBMCs were incubated at 4ºC with Fc-blocking reagent (Miltenyi Biotech, Germany) and stained for 30 minutes with the following monoclonal antibodies: anti-CD45 AF700, anti-CD14 APC-Cy7 and anti-CD19 PE-Cy7 (all three from Biolegend, CA, USA), anti-CD15 FITC and anti-CD3 AF67 (from Southern Biotech, USA), anti-CD16 AF647 and anti-CD34 PE (both from BD Biosciences Pharmingen, NJ, USA). After staining, erythrocytes from whole blood were lysed with Red Blood Cell Lysis Solution (Miltenyi Biotech, Germany). CD45^+^ CD15^+^ CD16^+^ neutrophils from whole blood and CD3^+^ T cells, CD19^+^ B cells, CD14^+^ monocytes, and CD34^+^ cells from PBMCs were FACS sorted in the BD FACSAriaII cell sorter (Beckton Dickinson, CA, USA) after excluding dead cells by DAPI staining.

After the COVID19 pandemic, as a consequence of restrictions in access to flow cytometry facilities, the leukocyte subpopulations were isolated from blood samples collected in EDTA- and heparin-containing tubes by using an immunomagnetic approach (EasySep, StemCell Technologies, USA), according to the manufacturer’s protocols. The purity of each obtained subpopulation was evaluated by flow cytometry.

***In vitro* studies of NLRP3-inflammasome activation**

HEK293T cells (CRL-11268, American Type Culture Collection) were maintained at 37ºC, 5% CO_2_ in a humidified incubator with Dulbecco’s modified Eagle’s medium (DMEM)/ F-12 media (Lonza, Basel, Switzerland) supplemented with 10% foetal calf serum (Life Technologies, CA, USA), 1% penicillin-streptomycin (Life Technologies, CA, USA) and 2 mM GlutaMAX (Life Technologies). Lipofectamine 2000 (Invitrogen, CA, USA) was used according to the manufacturer’s instructions for the transfection of HEK293T cells using 0.1 to 1 μg of pcDNA3.1 vector encoding for human NLRP3 wild-type (UniProt #Q96P20) or carrying the different variants as specified in Figure 5C. Intracellular ASC-speck formation was evaluated by seeding 7 x 10^5^ transfected HEK293T cells. MCC950 (10 μM, Sigma-Aldrich, MA, USA) was added to the cells 2h post-transfection. 24h post-transfection the ASC specks detection was performed by Time-of-Flight Inflammasome Evaluation (5) analysed by flow cytometry using LSRFortessa (BD Biosciences, NJ, USA) and the FCS express software (De Novo Software, CA, USA).

**Statistics**

Descriptive statistical methods were employed to summarize the clinical features, results of analytical tests and outcomes of administered treatments. Analyses of variables intergroups were performed using the McNemar test, the paired t-student test or the two-tailed unpaired t-test, as indicated. Differences were considered statistically significant at a p value of less than 0.05. All statistical analyses were performed using the Prism software version 10.3 (GraphPad Software, La Jolla, CA, USA).

**Supplementary Tables and Legends**

**Supplementary Table S1. Main Clinical Manifestations of Enrolled Patients**. Abbreviations: CAPS, cryopyrin-associated periodic syndromes; CINCA, chronic infantile, neurological, cutaneous and articular syndrome; NOMID, neonatal onset multisystem inflammatory disease; MWS, Muckle-Wells syndrome; FCAS, Familial cold-induced autoinflammatory syndrome; Oligo, Oligoarthritis; Poly, polyarthritis; n.a., not applicable.

|  | Phenotype | CAPS  Group | Disease  Onset  (years) | Trigger | Urticaria  Rash | Fever | Arthritis | Arthropathy | Conjuctivitis | Papilledema | Hearing  Loss | Aseptic  Meningitis | AA-Type  Amyloidosis |
| --- | --- | --- | --- | --- | --- | --- | --- | --- | --- | --- | --- | --- | --- |
| P1 | CINCA  NOMID |  | 16-20 | Cold | Yes | No | Yes  Oligo | Yes  Knees  Shoulders | No | Yes  Bilateral | Yes | Yes | No |
| P2 | CINCA  NOMID | Early  Onset | 0-5 | No | Yes | Yes | Yes  Oligo | Yes  Bilateral  Knees | Yes  Bilateral | No | Yes | Yes | No |
| P3 | MWS |  | 16-20 | No | Yes | Yes | Yes  Oligo | No | Yes  Bilateral | No | No | No | No |
| P4 | FCAS  MWS | Early  Onset | 0-5 | Cold | Yes | Yes | No | No | No | No | Yes  Bilateral | No | No |
| P5 | MWS | Late  Onset | 46-50 | Stress | Yes | Yes | No | No | No | Yes  Bilateral | Yes  Bilateral | No | Yes |
| P6 | Undefined  CAPS | Early  Onset | 0-5 | No | Yes | No | No | No | No | No | No | No | No |
| P7 | CINCA  NOMID | Early  Onset | 0-5 | No | Yes | Yes | Yes  Poly | Yes  Knees | No | Yes  Bilateral | No | Yes | No |
| P8 | Healthy |  | n.a. | n.a. | n.a. | n.a. | n.a. | n.a. | n.a. | n.a. | n.a. | n.a. | n.a. |
| P9 | MWS | Late  Onset | 51-55 | Stress | Yes | No | Yes  Poly | No | No | No | No | No | No |
| P10 | MWS |  | 11-15 | Trauma  Stress  Vaccines | Yes | Yes | No | No | No | No | No | No | No |
| P11 | MWS | Early  Onset | 6-10 | Cold  Infections | Yes | Yes | Yes | No | No | No | No | Yes | No |
| P12 | MWS | Early  Onset | 0-5 | Cold | Yes | No | Yes  Poly | No | No | No | Yes  Bilateral | No | No |
| P13 | MWS | Late  Onset | 76-80 | Infections  Trauma  Stress | Yes | Yes | Yes | No | Yes  Bilateral | No | No | Yes | Yes |
| P14 | MWS | Late  Onset | 61-65 | No | Yes | Yes | Yes  Poly | No | No | No | No | No | No |
| P15 | CINCA  NOMID | Early  Onset | 0-5 | Cold  Heat  Infections | Yes | Yes | Yes | Yes  Knees | No | No | No | No | No |
| P16 | MWS | Late  Onset | 56-60 | Heat  Stress | Yes | Yes | No | No | No | No | No | No | No |
| P17 | MWS | Late  Onset | 56-60 | No | Yes | Yes | Yes  Oligo | No | Yes  Bilateral | No | Yes  Bilateral | No | No |

**Supplementary Table S2. Results of Hematological Parameters of Enrolled Patients Before and During Treatment with anti-IL-1 Drugs.** Values inside each cell represent the mean and SD (brackets) of all collected data. Statistical analyses were performed using the two-tailed unpaired t-test. *p<0.05, **p<0.01, ***p<0.001, ****p<0.0001. Abbreviations: Hb, hemoglobin; MCV, mean corpuscular volume; RR, reference range; n.s., not significant.

|  | Hb  (g/L)  RR: 130-170 | | | MCV  (fL)  RR: 80-100 | | | Leukocyte count  (10^9^/L)  RR: 4.0-11.0 | | | Neutrophil count  (10^9^/L)  RR: 2-7 | | | Lymphocyte count  (10^9^/L)  RR: 0.9-4.5 | | | Platelets  (10^9^/L)  RR: 130-400 | | |
| --- | --- | --- | --- | --- | --- | --- | --- | --- | --- | --- | --- | --- | --- | --- | --- | --- | --- | --- |
|  | Before | During | p | Before | During | p | Before | During | p | Before | During | p | Before | During | p | Before | During | p |
| P1 | 108.8  (15.1) | 150.5  (14.4) | **** | 71.5  (7.2) | 84.8  (5.0) | ** | 16.1  (3.8) | 9.5  (1.4) | **** | 11.9  (3.9) | 6.2  (1.2) | **** | 2.5  (0.6) | 2.1  (0.5) | n.s. | 296  (57) | 203  (21) | **** |
| P2 | 110.8  (5.5) | 133.5  (8.9) | **** | 69.3  (1.2) | 80.2  (4.1) | **** | 22.1  (5.6) | 6.8  (1.5) | **** | 17.3  (6.3) | 4.3  (1.3) | **** | 4.2  (3.1) | 1.9  (0.5) | **** | 718  (100) | 304  (69) | **** |
| P3 | 148.5  (6.1) | 158.9  (6.1) | *** | 90.9  (2.0) | 93.3  (2.0) | ** | 14.5  (4.1) | 12.0  (2.6) | n.s. | 9.5  (4.4) | 6.7  (2.3) | * | 3.7  (0.6) | 3.7  (0.2) | n.s. | 233  (33) | 222  (53) | n.s. |
| P4 | 123.8  (5.9) | 136.2  (5.6) | *** | 84.1  (2.4) | 88.1  (2.8) | ** | 6.6  (2.7) | 4.1  (0.5) | * | 5.0  (2.7) | 2.6  (0.4) | * | 1.3  (0.2) | 1.2  (0.1) | n.s. | 247  (23) | 200  (13) | *** |
| P5 | 122.0  (5.0) | 116.6  (11.6) | n.s. | 85.3  (1.9) | 84.4  (3.9) | n.s. | 11.5  (1.7) | 10.9  (4.8) | n.s. | 6.6  (1.4) | 7.2  (4.4) | n.s. | 3.8  (1.1) | 2.3  (0.9) | *** | 407  (86) | 301  (52) | *** |
| P6 | 111  (0) | 125.8  (5.6) | n.s. | 76  (0) | 84.2  (2.2) | * | 14.5  (0) | 10.2  (1.3) | * | 7.1  (0) | 3.9  (0.7) | * | 5.7  (0) | 4.7  (1.1) | n.s. | 312  (0) | 268  (34) | n.s. |
| P7 | 105.4  (7.2) | 131.0  (8.5) | *** | 71.4  (2.0) | 87.9  (0.7) | **** | 18.6  (4.6) | 7.5  (0.6) | ** | 14.0  (5.5) | 4.6  (0.3) | * | 3.2  (1.2) | 1.9  (0.2) | n.s. | 652  (66) | 314  (38) | **** |
| P9 | 125.1  (3.2) | 139.7  (2.8) | **** | 93.4  (1.3) | 107.2  (3.2) | **** | 13.8  (4.3) | 4.5  (0.9) | **** | 10.9  (4.4) | 1.9  (0.7) | **** | 1.9  (0.4) | 1.9  (0.3) | n.s. | 678  (46) | 318  (90) | **** |
| P10 | 114.9  (5.3) | 136.8  (4.6) | **** | 82.3  (3.1) | 91.4  (2.5) | **** | 11.4  (3.2) | 7.2  (1.2) | **** | 9.2  (2.9) | 5.0  (1.1) | **** | 1.6  (0.3) | 1.5  (0.2) | n.s. | 446  (73) | 269  (19) | **** |
| P11 | 112.5  (3.5) | 119.1  (5.4) | n.s. | 95.4  (0.1) | 95.7  (2.8) | n.s. | 9.5  (2.8) | 7.2  (2.0) | n.s. | 7.3  (2.3) | 4.4  (1.9) | n.s. | 1.6  (0.3) | 1.9  (0.5) | n.s. | 315  (5) | 289  (34) | n.s. |
| P12 | 121.9  (15.4) | 144.7  (6.4) | **** | 87.9  (3.7) | 94.8  (1.1) | **** | 10.6  (2.1) | 6.5  (1.6) | **** | 7.9  (2.1) | 3.9  (1.6) | **** | 2.1  (0.3) | 1.8  (0.4) | n.s. | 422  (93) | 212  (24) | **** |
| P13 | 109.7  (8.2) | 127.3  (4.8) | **** | 80.9  (1.6) | 77.7  (2.4) | ** | 15.4  (3.0) | 6.2  (1.8) | **** | 11.9  (2.7) | 3.3  (1.3) | **** | 2.4  (1.0) | 1.8  (0.6) | n.s. | 394  (56) | 292  (44) | **** |
| P14 | 109.5  (6.2) | n.a. |  | 84.9  (2.4) | n.a. |  | 11.9  (1.9) | n.a. |  | 8.7  (2.4) | n.a. |  | 2.3  (0.7) | n.a. |  | 349  (50) | n.a. |  |
| P15 | 122.9  (10.2) | 130.8  (12.4) | n.s. | n.a. | n.a. |  | 10.5  (4.5) | 7.5  (1.4) | n.s. | 8.5  (4.3) | 5.5  (1.7) | n.s. | n.a. | n.a. |  | 344  (78) | 308  (64) | n.s. |
| P16 | 122.8  (6.9) | 138.7  (6.1) | ** | 88.8  (2.5) | 92.7  (0.5) | * | 14.9  (1.5) | 12.4  (2.0) | * | 8.2  (1.3) | 6.7  (1.8) | n.s. | 5.6  (1.4) | 4.4  (0.4) | n.s. | 416  (36) | 331  (17) | *** |
| P17 | 133.2  (9.5) | 147.5  (4.7) | ** | n.a. | n.a. |  | 22.2  (5.3) | 8.0  (0.8) | **** | 17.8  (5.5) | 3.3  (0.6) | **** | 3.0  (0.8) | 3.3  (0.2) | n.s. | 365  (55) | 155  (9) | **** |

**Supplementary Table S3. Results of Biochemical Parameters of Enrolled Patients Before and During Treatment with anti-IL-1 Drugs.** Values inside each cell represent the mean and SD (brackets) of all collected data. Statistical analyses were performed using the two-tailed unpaired t-test. *p<0.05, **p<0.01, ***p<0.001, ****p<0.0001. Abbreviations: ESR, erythrocyte sedimentation rate; n.a., data not available; n.s., not significant.

|  | C-reactive protein  (mg/L)  RR: <5 | | | ESR  (mm/h)  RR: <20 | | |
| --- | --- | --- | --- | --- | --- | --- |
|  | Before | During | p | Before | During | p |
| P1 | 68.6 (20.1) | 2.9 (2.6) | **** | 101.2 (17.8) | 19.8 (18.3) | **** |
| P2 | 38.3 (17.8) | 3.1 (2.2) | **** | 45.3 (5.8) | 11.8 (8.5) | **** |
| P3 | 37.3 (14.9) | 6.4 (5.9) | **** | 17.3 (6.0) | 5.9 (4.2) | **** |
| P4 | 40.9 (25.4) | 2.4 (2.5) | ** | 57.0 (7.2) | 15.7 (6.4) | **** |
| P5 | 29.7 (8.5) | 49.2 (94.7) | n.s. | 48.2 (30.0) | 43.2 (23.0) | n.s. |
| P6 | 7.9 (0) | 1.2 (1.4) | ** | 13 (0) | 2.1 (0.2) | **** |
| P7 | 37.0 (15.7) | 1.7 (0.6) | ** | 72.7 (12.5) | 16.0 (1.4) | ** |
| P9 | 37.8 (27.7) | 1.2 (0.8) | **** | 32.8 (18.2) | 10.6 (2.8) | ** |
| P10 | 39.5 (34.8) | 6.8 (5.5) | *** | 70.9 (32.2) | n.a |  |
| P11 | 30.5 (4.9) | 13.3 (12.2) | n.s. | 66.0 (12.7) | 27.5 (9.9) | ** |
| P12 | 69.8 (17.1) | 8.0 (5.4) | **** | 68.0 (36.2) | 19.1 (9.3) | **** |
| P13 | 38.7 (44.2) | 4.2 (3.1) | * | 59.1 (42.0) | 22.2 (15.5) | * |
| P14 | 32.1 (8.1) | n.a. |  | 93.0 (41.3) | n.a. |  |
| P15 | 56.7 (30.8) | 11.6 (19.8) | ** | 48.1 (22.4) | 34.0 (26.3) | n.s. |
| P16 | 56.7 (22.5) | 28.7 (13.2) | n.s. | 53.1 (20.7) | 35.3 (5.0) | n.s. |
| P17 | 100.1 (50.0) | 2.6 (2.5) | * | 40.8 (17.5) | 7.0 (5.3) | ** |

**Supplementary Table S4. Clinical Features of Patients with Early-onset CAPS versus Late-onset CAPS.** Statistical analyses were performed using Fisher’s exact test for binary variables, with no statistically significant differences observed. Disease onset differences were analyzed using the Mann-Whitney non-parametric test, which showed a significant p-value. Abbreviations: CAPS, cryopyrin-associated periodic syndromes; n.s., not significant.

|  | | **Early-onset CAPS (n: 6)** | **Late-onset CAPS (n: 6)** | **p** |
| --- | --- | --- | --- | --- |
| Disease Onset (Mean; range) (years) | | 1.13 (0.1-4) | 59 (47-76) | 0.005 |
| Triggering  Factors | Cold Exposure | 3 (50%) | 0 (0%) | 0.182 |
|  | Heat Exposure | 1 (16.7%) | 1 (16.7%) | 1 |
|  | Infections | 1 (16.7%) | 0 (0%) | 1 |
|  | Trauma | 0 (0%) | 0 (0%) | 1 |
|  | Psychological Stress | 0 (0%) | 4 (66.7%) | 0.061 |
|  | Vaccines | 0 (0%) | 0 (0%) | 1 |
| **Clinical Manifestations** | | | | |
| Recurrent Fever | | 4 (66.7%) | 5 (83.3%) | 1 |
| Gastrointestinal  Manifestations | Abdominal Pain | 0 (0%) | 2 (33.3%) | 0.455 |
|  | Nausea | 1 (16.7%) | 1 (16.7%) | 1 |
|  | Vomiting | 2 (33.3%) | 0 (0%) | 0.455 |
|  | Diarrhea | 0 (0%) | 1 (16.7%) | 1 |
|  | Constipation | 0 (0%) | 1 (16.7%) | 1 |
| Musculoskeletal  Manifestations | Arthralgia | 5 (83.3%) | 6 (100%) | 1 |
|  | Arthritis | 4 (66.7%) | 4 (66.7%) | 1 |
|  | Arthropathy | 3 (50%) | 0 (0%) | 0.182 |
|  | Myalgia | 2 (33.3%) | 5 (83.3%) | 0.242 |
|  | Myositis | 0 (0%) | 0 (0%) | 1 |
| Skin  Manifestations | Urticaria-like Lesiones | 6 (100%) | 6 (100%) | 1 |
|  | Erytemathous Papules/plaques | 3 (50%) | 4 (66.7%) | 1 |
|  | Purpuric Papules/plaques | 1 (16.7%) | 1 (16.7%) | 1 |
|  | Vesicular Lesions | 0 (0%) | 1 (16.7%) | 1 |
|  | Livedo Racemosa | 0 (0%) | 1 (16.7%) | 1 |
| Ocular  Manifestations | Conjuctivitis | 1 (16.7%) | 2 (33.3%) | 1 |
|  | Palpebral Edema | 0 (0%) | 1 (16.7%) | 1 |
|  | Episcleritis | 0 (0%) | 1 (16.7%) | 1 |
|  | Uveitis | 0 (0%) | 1 (16.7%) | 1 |
|  | Papilledema | 1 (16.7%) | 1 (16.7%) | 1 |
| Neurological  Manifestations | Neurosensorial Hearing Loss | 2 (33.3%) | 2 (33.3%) | 1 |
|  | Headache | 2 (33.3%) | 4 (66.7%) | 0.567 |
|  | Aseptic Meningitis | 2 (33.3%) | 1 (16.7%) | 1 |
| Others | Oral Ulcers | 2 (33.3%) | 0 (0%) | 0.455 |
|  | Genital Ulcers | 0 (0%) | 0 (0%) | 1 |
|  | Lymphadenopathies | 4 (66.7%) | 1 (16.7%) | 0.242 |
|  | Splenomegaly | 1 (16.7%) | 0 (0%) | 1 |
|  | Hepatomegaly | 2 (33.3%) | 1 (16.7%) | 1 |
|  | AA-Type Amyloidosis | 0 (0%) | 1 (16.7%) | 1 |
|  | Facial Dysmorphy | 3 (50%) | 0 (0%) | 0.182 |
|  | Frontal Bossing | 3 (50%) | 0 (0%) | 0.182 |
|  | Saddle Nose Deformity | 3 (50%) | 0 (0%) | 0.182 |

**Supplementary Table S5. Outcomes of Treatments in Enrolled Patients.** Abbreviations: NSAID, non-steroid anti-inflammatory drugs; IVIG, intravenous immunoglobulins; TNF, tumor necrosis factor; IL-6, interleukin 6; IL-1, interleukin 1.

| **Treatment** | **n (%)** | **Complete response** | **Partial response** | **Negative response** |
| --- | --- | --- | --- | --- |
| Antibiotics | 2/16 (12.5%) | 0 (0%) | 0 (0%) | 2 (100%) |
| Colchicine | 7/16 (43.75%) | 0 (0%) | 2 (28.6%) | 5 (71.4%) |
| NSAIDs | 12/16 (75%) | 0 (0%) | 4 (33.3%) | 8 (66.6%) |
| Glucocorticoids | 14/16 (87.5%) | 4 (28.6%) | 9 (64.3%) | 1 (7.1%) |
| Methotrexate | 5/16 (31.25%) | 0 (0%) | 1 (20%) | 4 (80%) |
| Mycophenolate mofetil | 2/16 (12.5%) | 0 (0%) | 0 (0%) | 2 (100%) |
| Azathioprine | 2/16 (12.5%) | 0 (0%) | 1 (50%) | 1 (50%) |
| Cyclosporine A | 4/16 (25%) | 0 (0%) | 1 (25%) | 3 (75%) |
| antihistamines | 7/16 (43.75%) | 0 (0%) | 0 (0%) | 7 (100%) |
| anti-IgE \| Omalizumab | 2/16 (12.5%) | 0 (0%) | 0 (0%) | 2 (100%) |
| anti-TNF \| etanercept | 2/16 (12.5%) | 0 (0%) | 1 (50%) | 1 (50%) |
| anti-TNF \| infliximab | 1/16 (6.25%) | 0 (0%) | 1 (100%) | 0 (0%) |
| anti-TNF \| adalimumab | 1/16 (6.25%) | 0 (0%) | 1 (100%) | 0 (0%) |
| anti-TNF \| golimumab | 1/16 (6.25%) | 0 (0%) | 1 (100%) | 0 (0%) |
| anti-IL-6 \| tocilizumab | 1/16 (6.25%) | 0 (0%) | 1 (100%) | 0 (0%) |
| anti-IL-1 \| anakinra | 14/16 (87.5%) | 13 (92.9%) | 1 (7.1%) | 0 (0%) |
| anti-IL-1 \| canakinumab | 8/16 (50.0%) | 8 (100%) | 0 (0%) | 0 (0%) |

**Supplementary Table S6. Classification of Post-zygotic *NLRP3* Variants**. ^1^Genome Build: GRCh38. ^2^RefSeq: NM_001243133.2. ^3^gnomAD version 4.0.0. ^4^CSVS version 5.0.0. ^5^ODINO project: Optimization of the diagnostic approach for inborn errors of immunity leading to hyper-inflammation | Project of the European Joint Programme on Rare Diseases - Joint Transnational Call 2022 (https://www.ejprarediseases.org/odino%ef%bf%bc/). ^6^Classification of pathogenicity of gene variants performed on the basis of standards and guidelines proposed in the consensus recommendations of the American College of Medical Genetics and Genomics and the Association for Molecular Pathology. ^7^ In patient 10, the post-zygotic variant is located at coordinate chr1:247424682; c.1233G>T. In the same triplet, the patient also carried at coordinate chr1:247424680 the c.1231C>T nucleotide transition (rs148478875). Abbreviations: Chr, chromosome; Ex, exon; gnomAD, Genome Aggregation Database; CSVS, Collaborative Spanish Variant Server; SIFT, Sorting Intolerant from Tolerant; CADD, Combined Annotation Dependent Depletion; ACMG / AMP, American College of Medical Genetics and Genomics / Association for Molecular Pathology; n.c., not classified; n.r., not registered; P, Pathogenic; LP, Likely Pathogenic; VUS, Variant of Uncertain Significance; Pro Dam, Probably Damaging; Pos Dam, Possibly Damaging; Ben, Benign; Del, Deleterious; Tol, Tolerated; Dis Caus, Disease Causing; Pol, Polymorphism; Germ, germline; Som, somatic; n.a., not available; n.t., not tested.

| Patient | Structural  Features | | | | Population  Genetics  Databases | | | Disease  Databases | | | Bioinformatic  Predictions | | | | Functional  Data | Reported Variant | ACMG / AMP Classification^6^ |
| --- | --- | --- | --- | --- | --- | --- | --- | --- | --- | --- | --- | --- | --- | --- | --- | --- | --- |
|  | Chromosome  position^1^ | Exon \| Intron | cDNA exchange^2^ | Predicted amino acid exchange | gnomAD (v4.1.0)^3^ | CSVS^4^ | ClinVar | | INFEVERS | Polyphen-2  (Hum Var Score) | | SIFT (Score) | Mutation Taster | CADD PHRED | ASC speck assay  (ODINO project)^5^ |  |  |
| P1 | chr1:247424356 | Ex 4 | c.907G>C | p.Asp303His | 0 | 0 | n.c. | | LP | Pro Dam  (1.000) | | Del  (0.02) | Dis Caus | 23.8 | Strongly  active | Yes  Som | P |
| P2 | chr1:247424356 | Ex 4 | c.907G>C | p.Asp303His | 0 | 0 | n.c. | | LP | Pro Dam  (1.000) | | Del  (0.02) | Dis Caus | 23.8 | Strongly  active | Yes  Som | P |
| P3 | chr1:247424357 | Ex 4 | c.908A>C | p.Asp303Ala | 0 | 0 | n.r. | | LP | Pro Dam  (0.999) | | Del  (0) | Dis Caus | 23.7 | Strongly  active | Yes  Som | LP |
| P4 | chr1:247424357 | Ex 4 | c.908A>G | p.Asp303Gly | 0 | 0 | P | | P | Pro Dam  (1.000) | | Del  (0) | Dis Caus | 23.9 | Strongly  active | Yes  Germ | LP |
| P5 | chr1:247424367 | Ex 4 | c.918A>T | p.Gln306His | 0 | 0 | n.r. | | n.r. | Pro Dam  (0.950) | | Tol  (0.08) | Pol | 10.42 | n.t. | Yes  Som | LP |
| P6 | chr1:247424369 | Ex 4 | c.920G>C | p.Gly307Ala | 0 | 0 | n.r. | | n.r. | Pos Dam  (0.517) | | Tol  (1.0) | Pol | 14.67 | n.t. | No | LP |
| P7 | chr1:247424369 | Ex 4 | c.920G>T | p.Gly307Val | 0 | 0 | P | | LP | Pos Dam  (0.636) | | Tol  (0.18) | Pol | 19.19 | Strongly  active | Yes  Germ | LP |
| P8 | chr1:247424492 | Ex 4 | c.1043C>T | p.Thr348Met | 0 | 0 | P | | P | Pro Dam  (0.955) | | Del  (0.01) | Dis Caus | 25.4 | Strongly  active | Yes  Germ &  Som | P |
| P9 | chr1:247424503 | Ex 4 | c.1054G>A | p.Ala352Thr | 0 | 0 | LP | | LP | Pro Dam  (0.946) | | Del  (0.04) | Dis Caus | 25.7 | Strongly  active | Yes  Som | LP |
| P10 ^7^ | chr1:247424682 | Ex 4 | c.1233G>T | p.Leu411Phe | 0 | 0 | n.r. | | LP | Ben  (0.322) | | Del  (0.005) | Dis Caus | n.a. | Strongly  active | Yes  Som | LP |
| P11 | chr1:247424756 | Ex 4 | c.1307C>T | p.Thr436Ile | 0 | 0 | P  LP | | P | Pro Dam  (0.997) | | Del  (0) | Dis Caus | 23.9 | Strongly  active | Yes  Germ | LP |
| P12 | chr1:247425018 | Ex 4 | c.1569C>A | p.Phe523Leu | 0 | 0 | n.c. | | LP | Pos Dam  (0.510) | | Del  (0) | Dis Caus | 24.3 | Strongly  active | Yes  Germ &  Som | LP |
| P13 | chr1:247425137 | Ex 4 | c.1688A>G | p.Tyr563Cys | 0 | 0 | n.r. | | n.c. | Ben  (0.068) | | Tol  (0.08) | Dis Caus | 22.5 | Strongly  active | Yes  Som | LP |
| P14 | chr1:247425147 | Ex 4 | c.1698C>G | p.Phe566Leu | 0 | 0 | n.r. | | n.r. | Ben  (0.093) | | Tol  (0.32) | Pol | 12.66 | Strongly  active | Yes  Germ &  Som | LP |
| P15 | chr1:247425148 | Ex 4 | c.1699G>A | p.Glu567Lys | 0 | 0 | P  LP | | LP | Ben  (0.056) | | Tol  (0.05) | Dis Caus | 22.4 | Strongly  active | Yes  Germ &  Som | LP |
| P16 | chr1:247425155 | Ex 4 | c.1706G>A | p.Gly569Glu | 0 | 0 | VUS | | n.r. | Ben  (0.186) | | Tol  (0.12) | Dis Caus | 22.1 | n.t. | No | LP |
| P17 | chr1:247425355 | Ex 4 | c.1906C>G | p.Gln636Glu | 0.00032% | 0 | n.r. | | n.c. | Pos Dam  (0.542) | | Del  (0) | Dis Caus | 23.7 | Strongly  active | Yes  Som | LP |

**Supplementary Table S7. Clonal Dynamics of *NLRP3* Mosaicism Over Time**. ^1^RefSeq: NM_001243133.2. ^2^Values are the mean (SD) of three to six independent experiments, expressed as percentage. ^3^Defined as the number of months between the first analyzed sample (time 0) and the subsequent blood collections. Abbreviations: VAF, variant allele frequency; Cov, coverage.

| Patient | *NLRP3* Exchange^1^ | 1^st^ Sample  Peripheral Blood  (Genetic Diagnosis) | | | 2^nd^ Sample  Peripheral Blood | | | | 3^rd^ Sample  Peripheral Blood | | | 4^th^ Sample  Peripheral Blood | | |
| --- | --- | --- | --- | --- | --- | --- | --- | --- | --- | --- | --- | --- | --- | --- |
|  |  | VAF^2^ | Cov^2^ | Months^3^ | | VAF^2^ | Cov^2^ | Months^3^ | | VAF^2^ | Cov^2^ | Months^3^ | VAF^2^ | Cov^2^ |
| P1 | c.907G>C | 14.10%  (0.9) | 9988x  (1746) | 38 | | 12.1%  (0.5) | 2635x  (21) | 143 | | 7.9%  (0.2) | 11499x  (1315) |  |  |  |
| P2 | c.907G>C | 17.1%  (1.3) | 37091x  (20804) | 209 | | 10.0%  (0.9) | 3888x  (156) |  | |  |  |  |  |  |
| P3 | c.908A>C | 34.8%  (0.8) | 31073x  (7565) |  | |  |  |  | |  |  |  |  |  |
| P4 | c.908A>G | 17.2%  (1.9) | 1756x  (381) | 62 | | 15.8%  (0.2) | 7499x  (725) |  | |  |  |  |  |  |
| P5 | c.918A>T | 4.9%  (0.3) | 11083x  (4384) | 2 | | 4.9%  (0.3) | 9212x  (693) |  | |  |  |  |  |  |
| P6 | c.920G>C | 2.4%  (0.3) | 22111x  (3488) | 19 | | 2.4%  (0.4) | 16442x  (797) |  | |  |  |  |  |  |
| P7 | c.920G>T | 9.4%  (0.5) | 28048x  (5221) | 152 | | 6.4%  (0.4) | 16905x  (1454) | 259 | | 7.4%  (0.4) | 1851x  (434) |  |  |  |
| P8 | c.1043C>T | 3.1%  (0.2) | 24063x  (3876) | 104 | | 3.0%  (0.0) | 29972x  (9733) | 244 | | 2.4%  (0.3) | 9413x  (1284) |  |  |  |
| P9 | c.1054G>A | 22.7%  (0.5) | 9449x  (3019) | 88 | | 2.2%  (0.1) | 6679x  (876) |  | |  |  |  |  |  |
| P10 | c.1233G>T | 16.4%  (0.6) | 16987x  (5664) | 82 | | 24.3%  (0.7) | 17868x  (1325) | 255 | | 33.0%  (1.4) | 12322x  (1953) |  |  |  |
| P11 | c.1307C>T | 9.0%  (0.5) | 3043x  (1658) | 3 | | 9.0%  (1.2) | 2735x  (1440) | 18 | | 9.5%  (0.8) | 3042x  (1610) | 55 | 12.0%  (0.2) | 3442x  (30) |
| P12 | c.1569C>A | 10.1%  (0.6) | 21788x  (2640) | 155 | | 8.6%  (0.3) | 5084x  (1108) |  | |  |  |  |  |  |
| P13 | c.1688A>G | 8.0%  (1.2) | 4187x  (1881) | 104 | | 13.5%  (0.6) | 5745x  (911) |  | |  |  |  |  |  |
| P14 | c.1698C>G | 1.3%  (0.1) | 10818x  (2311) | 81 | | 1.7%  (0.0) | 10955x  (1388) | 89 | | 1.6%  (0.1) | 14058x  (2226) | 93 | 1.5%  (0.1) | 18807x  (4512) |
| P15 | c.1699G>A | 2.4%  (0.2) | 3474x  (877) | 8 | | 2.8%  (0.1) | 7018x  (1762) |  | |  |  |  |  |  |
| P16 | c.1706G>A | 1.3%  (0.1) | 18395x  (308) | 3 | | 1.1%  (0.2) | 12849x  (3510) | 8 | | 1.5%  (0.3) | 7725x  (1140) | 30 | 2.2%  (0.2) | 13270x  (1057) |
| P17 | c.1906C>G | 18.0%  (0.7) | 4446x  (309) | 4 | | 23.4%  (2.6) | 3510x  (666) | 6 | | 10.4%  (0.2) | 7031x  (204) |  |  |  |

**Supplementary Table S8. Distribution of Mosaicism**. ^1^RefSeq: NM_001243133.2. ^2^Defined as CD45^+^ CD15^+^ CD16^+^. ^3^Defined as CD45^+^ CD14^+^. ^4^Defined as CD45^+^ CD3^+^. ^5^Defined as CD45^+^ CD19^+^. ^6^Determined by flow cytometry analyses after cell isolation. ^7^Values are the mean (SD) of three independent experiments, expressed as percentage. ^8^Mutant allele frequency was not investigated in monocytes and B cells due to low purity of isolated cells. Abbreviations: VAF, variant allele frequency; Cov, coverage.

| Patient | cDNA  *NLRP3*  Exchange^1^ | Neutrophils^2^ | | | | Monocytes^3^ | | | | T cells^4^ | | | | B cells^5^ | | | | Nails | |
| --- | --- | --- | --- | --- | --- | --- | --- | --- | --- | --- | --- | --- | --- | --- | --- | --- | --- | --- | --- |
|  |  | Purity^6^ | VAF^7^ | Cov^7^ | Purity^6^ | | VAF^7^ | Cov^7^ | Purity^6^ | | VAF^7^ | Cov^7^ | Purity^6^ | | VAF^7^ | Cov^7^ | VAF^7^ | | Cov^7^ |
| P1 | c.907G>C | 98.8% | 8.0%  (0.5) | 9153x  (1628) | 96.2% | | 6.6%  (0.4) | 8868x  (539) | 98.6% | | 11.4%  (0.4) | 10190x  (551) | 98.6% | | 8.4%  (0.2) | 5745x  (4416) | 12.4%  (0.1) | | 5017x  (2362) |
| P2 | c.907G>C | 96.2% | 7.8%  (1.2) | 5235x  (224) | 91.7% | | 7.1%  (0.6) | 4473x  (351) | n.a. | | 18.2%  (0.6) | 24489x  (1478) | n.a. | | 14.9%  (3.6) | 20785x  (8253) | n.d. | | n.d. |
| P4 | c.908A>G | 88.0% | 14.7%  (0.3) | 6189x  (640) | 91.3% | | 15.6%  (2.2) | 4312x  (1659) | 99.0% | | 17.7%  (1.2) | 5145x  (3454) | 99.5% | | 16.1%  (1.4) | 3980x  (1283) | 13.4%  (0.9) | | 4129x  (1697) |
| P5^8^ | c.918A>T | n.a. | 6.0%  (0.5) | 8652x  (3056) | n.a. | | n.p. | n.p. | n.a. | | 1.1%  (0.1) | 10039x  (2064) | n.a. | | n.p. | n.p. | 0.1%  (0.1) | | 14253x  (12668) |
| P8 | c.1043C>T | 91.9% | 1.7%  (0.1) | 10142x  (624) | 95.2% | | 1.6%  (0.1) | 7151x  (1302) | 97.2% | | 4.3%  (0.3) | 9555x  (1888) | 97.4% | | 2.8%  (0.7) | 5388x  (1401) | 1.1%  (1.2) | | 3897x  (580) |
| P9 | c.1054G>A | 88.5% | 3.1%  (0.2) | 6077x  (1256) | 95.3% | | 2.8%  (0.4) | 4128x  (1138) | 93.5% | | 0.6%  (0.1) | 5900x  (831) | 97.6% | | 2.1%  (0.6) | 5536x  (60) | 0.0%  (0) | | 5712x  (453) |
| P10 | c.1233G>T | 91.1% | 36.2%  (0.3) | 13605x  (2257) | 98.3% | | 33.4%  (1.2) | 6147x  (3524) | 97.9% | | 15.5%  (1.1) | 9073x  (3012) | 98.7% | | 23.8%  (2.0) | 2988x  (1528) | 4.3%  (0.8) | | 4063x  (2061) |
| P11 | c.1307C>T | 96.6% | 9.8%  (0.3) | 3330x  (1048) | 98.4% | | 10.7%  (1.7) | 3249x  (2943) | 99.4% | | 3.7%  (0.4) | 5753x  (794) | 99.2% | | 10.8%  (0.9) | 1046x  (257) | 4.8%  (0.9) | | 3728x  (1972) |
| P12 | c.1569C>A | 94.9% | 8.7%  (0.4) | 4397x  (226) | 96.5% | | 9.8%  (2.4) | 6095x  (7107) | 96.1% | | 3.9%  (0.1) | 2752x  (964) | 96.1% | | 9.2%  (1.8) | 1486x  (1970) | 4.2%  (1.2) | | 1399x  (690) |
| P13 | c.1688A>G | 96.9% | 22.3%  (1.6) | 3174x  (651) | 95.7% | | 17.3%  (0.7) | 3190x  (1284) | 97.5% | | 0.7%  (0.1) | 6047x  640) | 98.6% | | 3.3%  (0.6) | 1467x  (806) | 1.1%  (0.1) | | 6030x  (595) |
| P14 | c.1698C>G | 98.1% | 2.1%  (0.0) | 14148x  (2591) | 93.4% | | 2.4%  (0.1) | 13980x  (1796) | 96.6% | | 0.3%  (0.1) | 14229x  (1130) | 99.6% | | 3.9%  (0.1) | 14168x  (2648) | 1.1%  (0.7) | | 11840x  (1778) |
| P15 | c.1699G>A | 95.3% | 4.4%  (0.5) | 9370x  (3215) | 97.3% | | 3.8%  (1.0) | 7035x  (2119) | 96.8% | | 4.2%  (0.2) | 7350x  (16) | 98.3% | | 4.1%  (1.2) | 6609x  (3027) | n.d. | | n.d. |
| P16 | c.1706G>A | 90.4% | 2.1%  (0.3) | 8583x  (1067) | 89.9% | | 2.6%  (0.1) | 8328x  (1333) | 93.2% | | 0.1%  (0.0) | 8612x  (484) | 93.4% | | 0.6%  (0.1) | 8831x  (1208) | 0.2%  (0.2) | | 8215x  (404) |
| P17 | c.1906C>G | n.d. | 24.6%  (3.0) | 3197x  (365) | n.d. | | 30.4%  (2.7) | 3642x  (278) | n.d. | | 0.0%  (0) | 3002x  (529) | n.d. | | 0.0%  (0) | 3097x  (662) | n.d. | | n.d. |

**Supplementary Table S9. List of analyzed genes associated with autoinflammatory diseases.** Abbreviations: ADA2, adenosin deaminase 2; ARPC1B, actin related protein C1B; NOCARH, Neonatal-onset cytopenia, autoinflammation, rash and hemophagocytic lymphohistiocytosis; IL-10, interleukin-10; DIRA, deficiency of interleukin-1 receptor antagonist; DITRA, deficiency of interleukin-36 receptor antagonist; FMF, familial Mediterranean Fever; PAAND, Pyrin-associated autoinflammation with neutrophilic dermatosis; MK, mevalonate kinase; AIFEC, Autoinflammation with infantile enterocolitis; NAIAD, NLRP1-associated autoinflammation with arthritis and dyskeratosis; FCAS2, familial cold-induced autoinflammatory syndrome type 2; CAPS, cryopyrin-associated periodic syndromes; PLAID, PLCG2-associated antibody deficiency, and immune dysregulation; APLAID, Autoinflammation and PLCG2-associated antibody deficiency, and immune dysregulation; CANDLE, Chronic atypical neutrophilic dermatosis with lipodystrophy and elevated temperature; PRAAS, Proteasome-associated autoinflammatory syndrome; PAPA, Pyogenic arthritis, pyoderma gangrenosum and acne; Hz/Hc, Hyperzincemia and hypercalprotectinemia syndrome; HOIL-1, Heme-oxidized IRP2 ubiquitin ligase 1; CRIA, cleavage-resistant RIPK1-induced autoinflammatory syndrome; HOIP, HOIL-1-interacting protein; SAVI, STING-associated vasculopathy with onset in infancy; TRAPS11, TNFRSF11A-associated periodic syndrome; TRAPS, TNF Receptor I-associated periodic syndrome; SIFD, Sideroblastic anemia, B-cell immunodeficiency, periodic fevers, and developmental delay; PFIT, periodic fever, immunodeficiency, and thrombocytopenia syndrome.

| **Gene** | **Disease** | **Ref Seq** |  | **Gene** | **Disease** | **Ref Seq** |
| --- | --- | --- | --- | --- | --- | --- |
| *ADA2* | ADA2 Deficiency | NM_001282225.1 |  | *PLCG2* | PLAID-APLAID | NM_002661.3 |
| *ADAR* | Aicardi-Goutières syndrome Type 6 | NM_001111.5 |  | *POMP* | CANDLE/PRAAS | NM_015932.5 |
| *AP1S3* | Pustular Psoriasis | NM_001039569.1 |  | *PSMA3* | CANDLE/PRAAS | NM_002788.3 |
| *ARPC1B* | ARPC1B Deficiency | NM_005720.4 |  | *PSMB4* | CANDLE/PRAAS | NM_002796.2 |
| *CARD14* | Pustular Psoriasis | NM_024110.4 |  | *PSMB8* | CANDLE/PRAAS | NM_148919.3 |
| *CDC42* | NOCARH | NM_001791.4 |  | *PSMB9* | CANDLE/PRAAS | NM_002800.4 |
| *IFIH1* | Aicardi-Goutières syndrome Type 7 | NM_022168.3 |  | *PSMG2* | CANDLE/PRAAS | NM_020232.4 |
| *IL10* | IL-10 Deficiency | NM_000572.2 |  | *PSTPIP1* | PAPA-Hz/Hc | NM_003978.3 |
| *IL10RA* | IL-10R1 Deficiency | NM_001558.3 |  | *RBCK1* | HOIL-1 Deficiency | NM_031229.3 |
| *IL10RB* | IL-10R2 Deficiency | NM_000628.4 |  | *RELA* | Rel-A Haploinsufficiency | NM_021975.3 |
| *IL1RN* | DIRA | NM_173842.2 |  | *RIPK1* | CRIA | NM_003804.5 |
| *IL36RN* | DITRA | NM_173170.1 |  | *RNASEH2A* | Aicardi-Goutières syndrome Type 4 | NM_006397.2 |
| *LACC1* | Lacasse Deficiency | NM_153218.3 |  | *RNASEH2B* | Aicardi-Goutières syndrome Type 2 | NM_024570.3 |
| *LPIN2* | Majeed syndrome | NM_014646.2 |  | *RNASEH2C* | Aicardi-Goutières syndrome Type 3 | NM_032193.3 |
| *MEFV* | FMF-PAAND | NM_000243.2 |  | *RNF31* | HOIP Deficiency | NM_017999.4 |
| *MVK* | MK Deficiencies | NM_000431.3 |  | *SAMHD1* | Aicardi-Goutières syndrome Type 5 | NM_015474.3 |
| *NCSTN* | Hidradenitis suppurativa | NM_015331.2 |  | *TMEM173* | SAVI | NM_198282.3 |
| *NLRC4* | AIFEC | NM_021209.4 |  | *TNFAIP3* | A20 Haploinsufficiency | NM_006290.3 |
| *NLRP1* | NAIAD | NM_033004.3 |  | *TNFRSF11A* | TRAPS11 | NM_003839.3 |
| *NLRP12* | FCAS2 | NM_144687.2 |  | *TNFRSF1A* | TRAPS | NM_001065.3 |
| *NLRP3* | CAPS | NM_001243133.2 |  | *TREX1* | Aicardi-Goutières syndrome Type 1 | NM_033629.5 |
| *NOD2* | Blau syndrome | NM_022162.2 |  | *TRNT1* | SIFD | NM_182916.2 |
| *OTULIN* | Otulipenia | NM_138348.5 |  | *WDR1* | PFIT | NM_017491.4 |

**Legends of Supplementary Figures**

**Supplementary Figure S1. Sanger chromatograms showing clonal dynamics of *NLRP3* mosaicism in P10.** The gray arrows indicate the position where the postzygotic nucleotide variant is located, whereas green arrows indicate the position of a common nucleotide substitution (rs148478875). The numbers above each panel, preceded by the letter M, indicate the months that separate the analyzed sample from the baseline sample (M0). Percentages highlighted in red indicate the mean variant allele frequency obtained in three independent experiments of amplicon-based deep sequencing.

**Supplementary Figure S2. Sanger chromatograms showing clonal dynamics of *NLRP3* mosaicism in P13.** The gray arrows indicate the position where the postzygotic nucleotide variant is located. The numbers above each panel, preceded by the letter M, indicate the months that separate the analyzed sample from the baseline sample (M0). Percentages highlighted in red indicate the mean variant allele frequency obtained in three independent experiments of amplicon-based deep sequencing.

**Supplementary Figure S3. Sanger chromatograms showing clonal dynamics of *NLRP3* mosaicism in P9.** The gray arrows indicate the position where the postzygotic nucleotide variant is located. The numbers above each panel, preceded by the letter M, indicate the months that separate the analyzed sample from the baseline sample (M0). Percentages highlighted in red indicate the mean variant allele frequency obtained in three independent experiments of amplicon-based deep sequencing.

**Supplementary Figure S4. Sanger chromatograms showing clonal dynamics of *NLRP3* mosaicism in P1.** The gray arrows indicate the position where the postzygotic nucleotide variant is located. The numbers above each panel, preceded by the letter M, indicate the months that separate the analyzed sample from the baseline sample (M0). Percentages highlighted in red indicate the mean variant allele frequency obtained in three independent experiments of amplicon-based deep sequencing.

**Supplementary Figure S5. Sanger chromatograms showing clonal dynamics of *NLRP3* mosaicism in P2.** The gray arrows indicate the position where the postzygotic nucleotide variant is located. The numbers above each panel, preceded by the letter M, indicate the months that separate the analyzed sample from the baseline sample (M0). Percentages highlighted in red indicate the mean variant allele frequency obtained in three independent experiments of amplicon-based deep sequencing.
