## Supplementary figures and images for "Clinical Features, Outcomes of Treatments, Inflammasome Function and Longitudinal Clonal Dynamics into *NLRP3* Mosaicism: Evidence from the Largest Cryopyrin-associated Periodic Syndromes Cohort to Date"

### Supplementary Figure S01

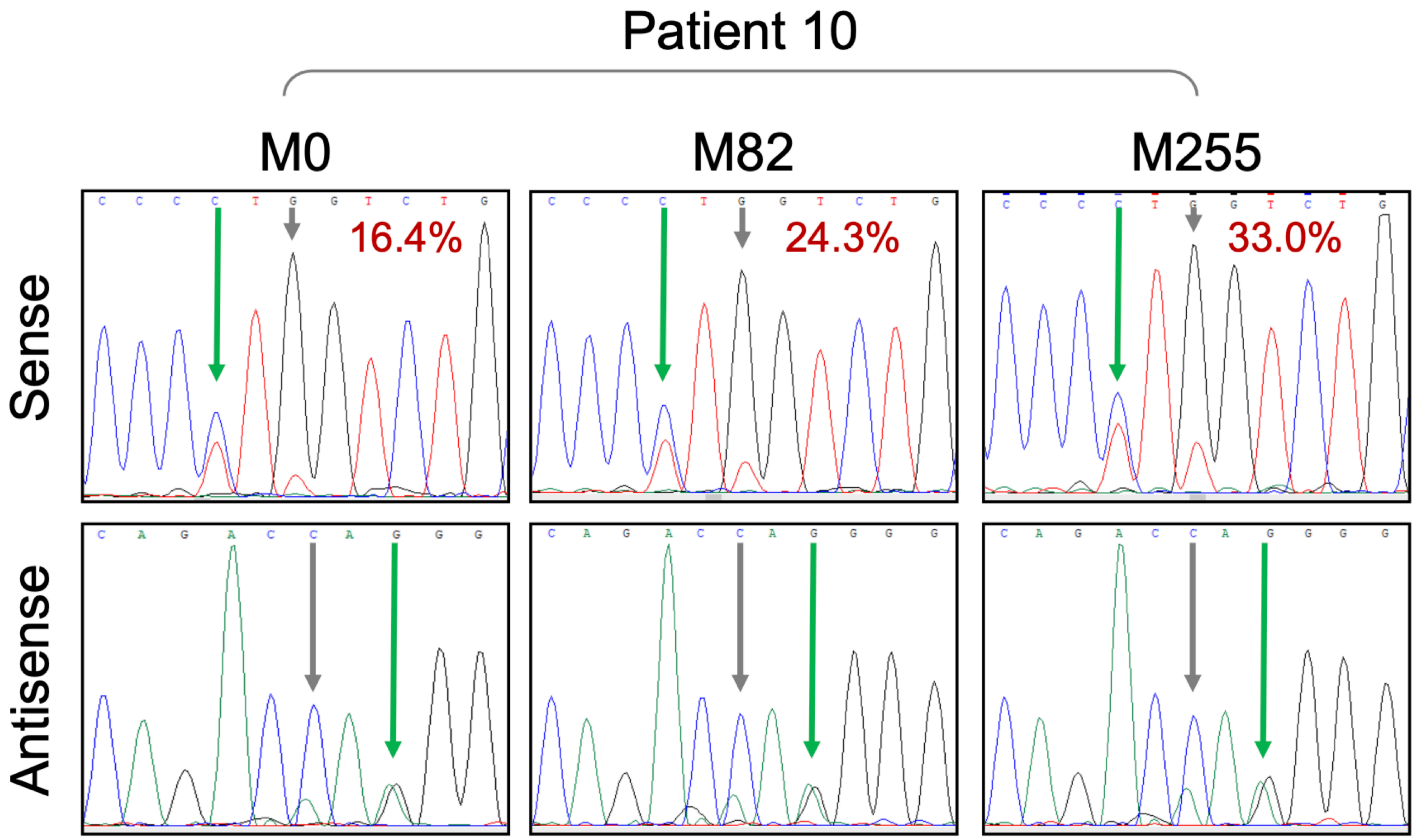

### Supplementary Figure S02

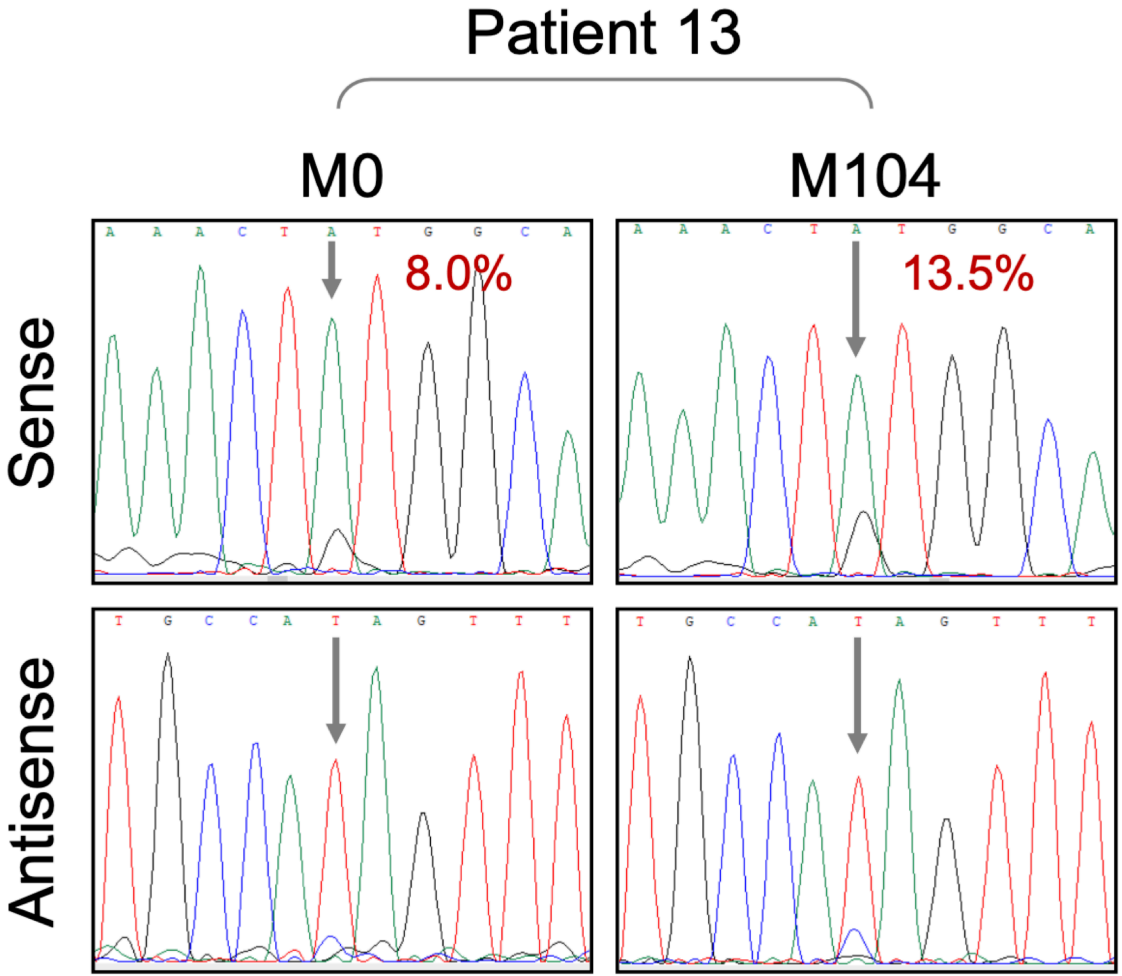

### Supplementary Figure S03

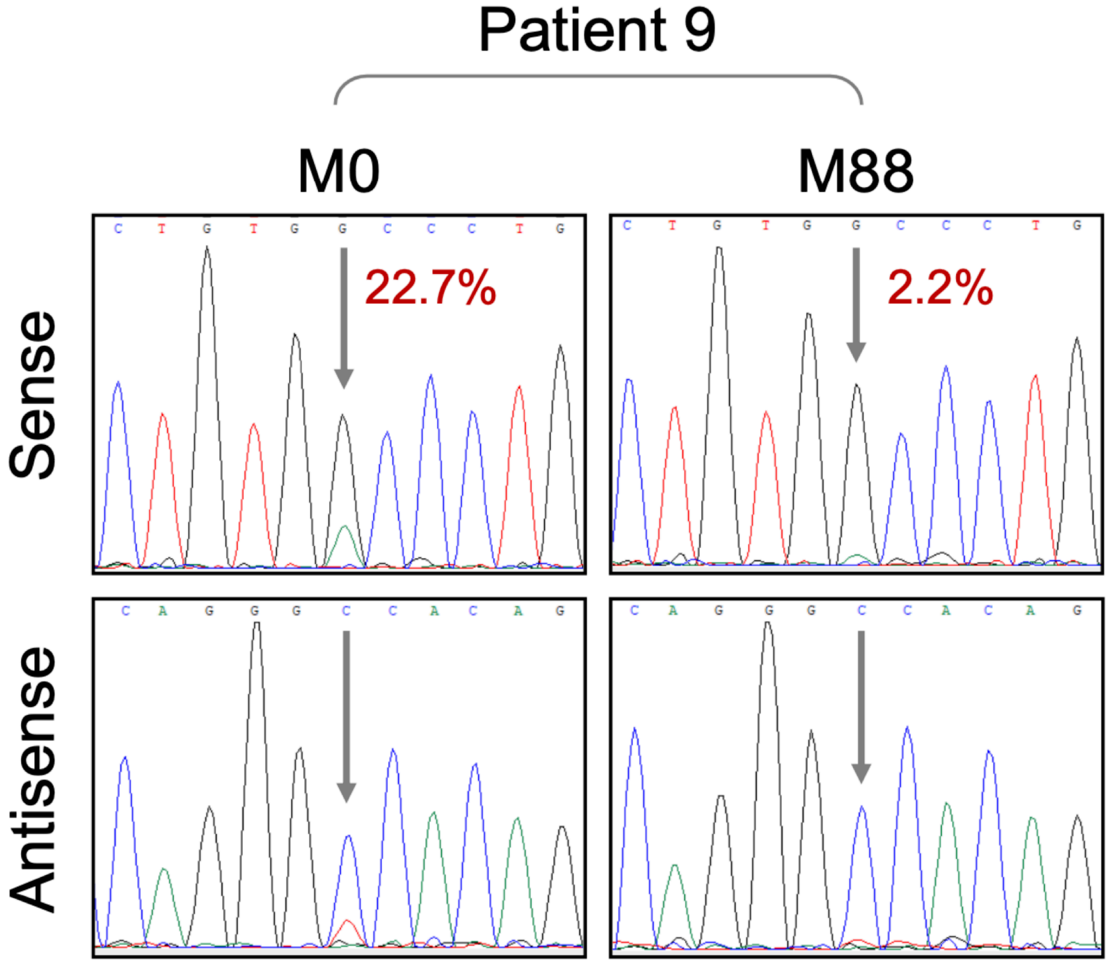

### Supplementary Figure S04

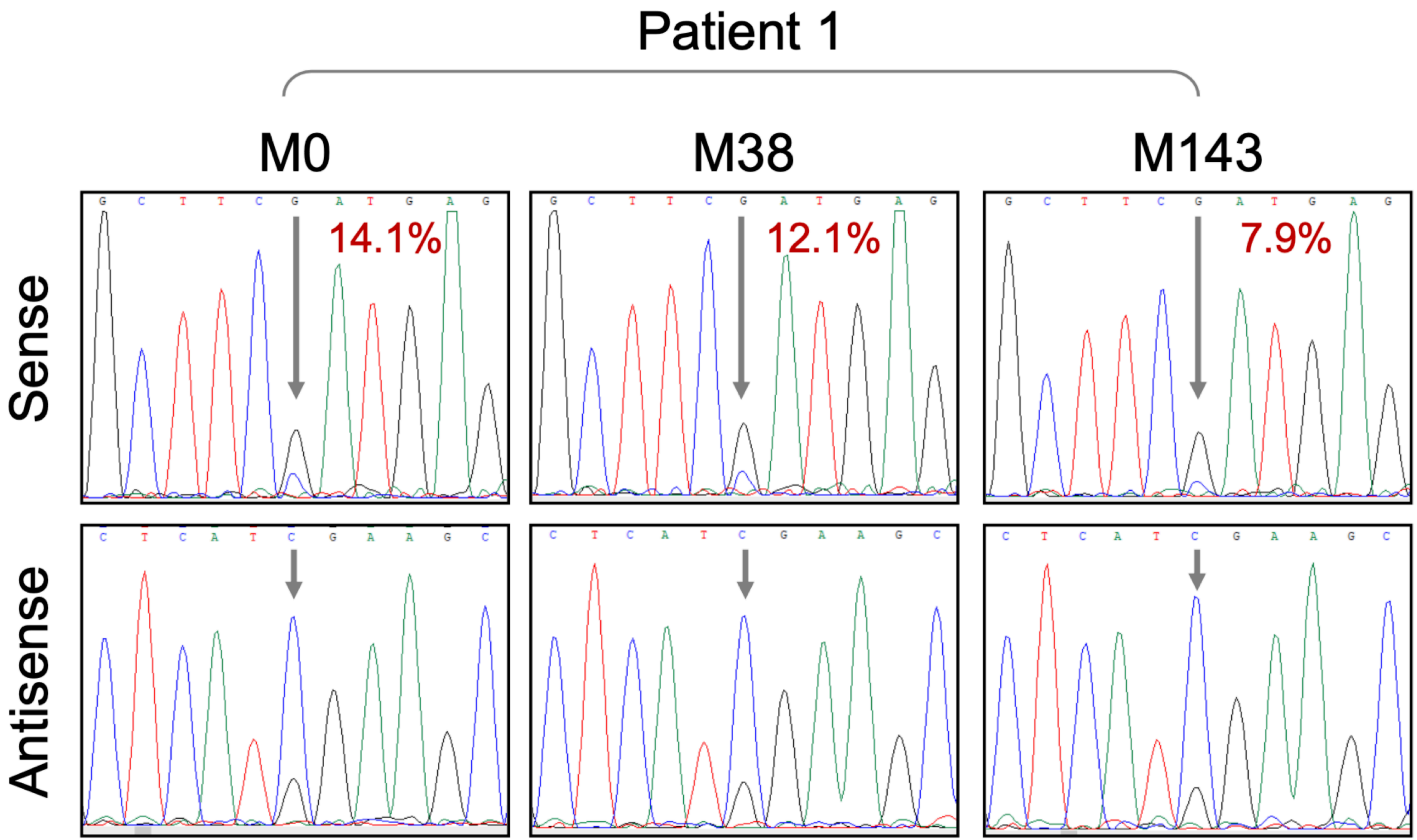

### Supplementary Figure S05

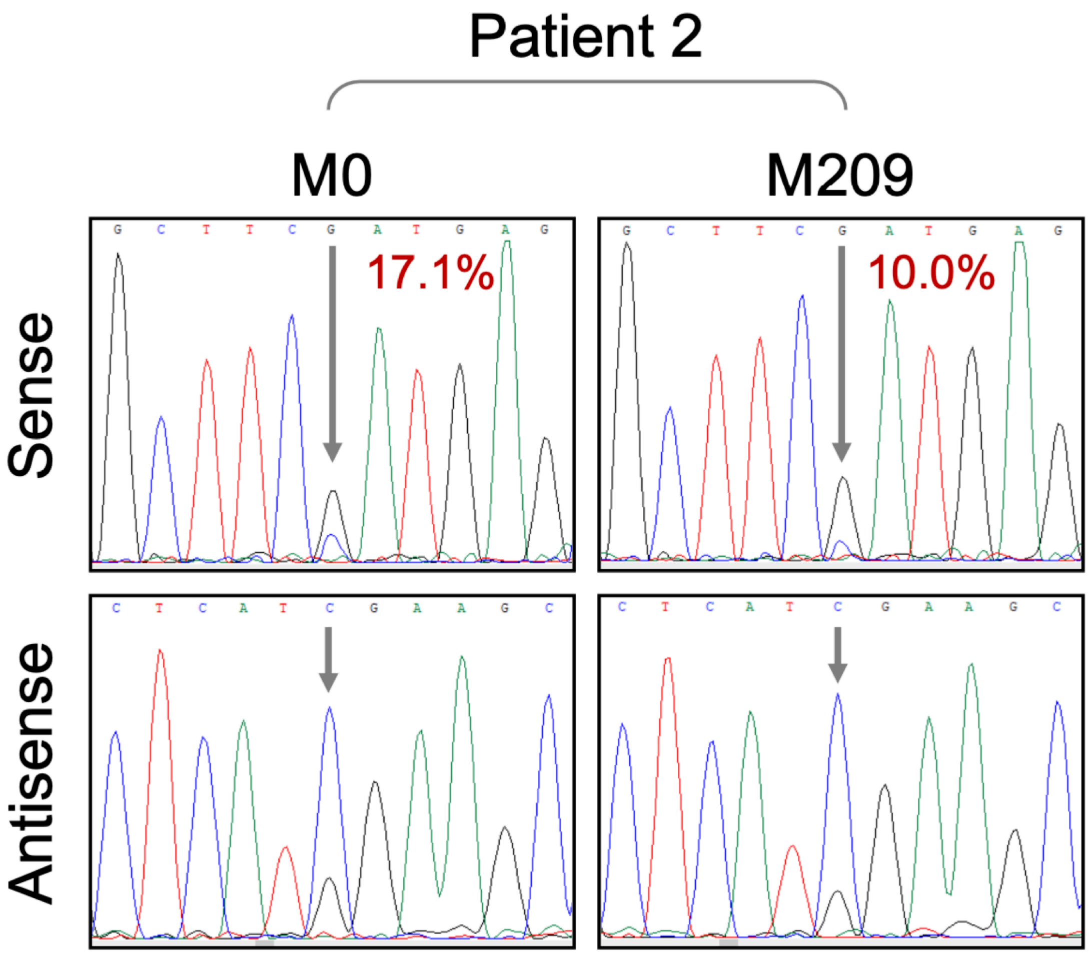
